# A systematic review investigating emerging trends between Extreme Weather Events (EWEs) and infectious disease outbreaks in South Africa

**DOI:** 10.1101/2025.03.06.25323483

**Authors:** Natalie Dickinson, Llinos Haf Spencer, Caroline Miller, Nisha Nadesanreddy, Serestina Viriri, Muhammad Zeeshan Shakir, Michael Gebreslasie, David Ndzi, Ozayr Harron Mahomed, Saloshni Naidoo, Mary Lynch, Fiona L. Henriquez

**Affiliations:** School of Health and Life Sciences, University of the West of Scotland, Paisley, UK; Faculty of Nursing and Midwifery, Royal College of Surgeons in Ireland, Dublin, Ireland; Faculty of Life Sciences and Education, University of South Wales, Cardiff, UK; Discipline of Public Health, Howard College, University of Kwazulu-Natal, Durban, South Africa; School of Mathematics, Statistics, and Computer Science, University of Kwazulu-Natal, Durban, South Africa; School of Computing, Engineering and physical Sciences, University of the West of Scotland, Paisley, UK; School of Electrical and Mechanical Engineering, University of Portsmouth, UK; Department of Civil and Environmental Engineering, University of Strathclyde, Glasgow, UK

**Keywords:** early warning system, extreme weather events, South Africa, floods, drought, health system resilience, community health

## Abstract

Extreme weather events (EWEs) are increasing in frequency and intensity due to climate change, exacerbating health risks, particularly for vulnerable populations. Infectious diseases are climate-sensitive, yet microbial dynamics during and after EWEs remain poorly understood. This systematic review examines emerging trends in infectious disease outbreaks following EWEs in South Africa.

A comprehensive search across fifteen electronic databases was conducted using Cochrane systematic review principles. Studies focusing on infectious diseases related to EWEs in South Africa were included, considering all study designs. A PRISMA diagram details the screening process, and quality appraisal was conducted using JBI checklist tools or a mixed-methods assessment tool.

No primary studies specifically linking recent infectious disease outbreaks to EWEs in South Africa were identified. Existing literature focuses on symptom presentation rather than microbial identification. However, findings indicate an increase in infectious diseases post-EWEs, with three key themes emerging: (1) climatic impacts on infectious diseases, (2) population vulnerabilities, and (3) the role of infrastructure and policy. The review highlights the need for longitudinal microbiological data to improve outbreak prediction and preparedness.

This review underscores a significant research gap and calls for an integrated approach combining environmental monitoring with pathogen diagnostics. Large-scale longitudinal studies and enhanced collaboration between education and healthcare sectors are needed. Early Warning Systems should incorporate climate variables to predict disease outbreaks, and a One Health approach is essential to address climate-driven infectious disease risks effectively.

## Background

The surge in EWEs in the past 25 years has resulted in the most devastating impacts; in the period 2000-2019, six of the ten countries most affected by EWEs were on the African continent (Centre for Research on the Epidemiology of Disasters (CRED), 2019). In South Africa, the frequency and intensity of extreme weather events (EWEs), including droughts, floods, and heatwaves, have increased due to climate change (Calvin et al., 2023) significantly influencing public health, particularly the spread of infectious diseases (Haines et al., 2006; McMichael, 2015; Saatchi et al., 2024).

The country’s diverse climate zones, coupled with socio-economic disparities, multidimensional inequalities and limited healthcare infrastructure in certain regions create an environment where climate-induced disease outbreaks pose a critical challenge (Hassell et al., 2017; Liao et al., 2024; Meurens et al., 2021; Moyo et al., 2023; World Organisation for Animal Health, 2015). Infections, such as malaria, cholera, and diarrheal diseases, are closely linked to environmental changes, exacerbated by heavy rainfall, flooding, and droughts (McMichael, 2015). On the continent of Asia, flood-related cholera outbreaks are commonplace, particularly in regions with poor sanitation and overcrowding (Shackleton et al., 2024). In Pakistan, extreme monsoon floods in 2022, led to a significant rise in waterborne diseases, including hepatitis E and acute gastroenteritis, as millions were displaced and forced to consume contaminated water (Raza et al., 2023). Peru and Brazil have witnessed a resurgence of vector- and waterborne diseases, including dengue fever and leptospirosis, in the wake of El Niño-driven flooding events (Acosta-España et al., 2024). The Amazon region is particularly vulnerable, as rising temperatures and prolonged wet conditions create ideal breeding grounds for mosquitoes, intensifying malaria transmission (MacDonald & Mordecai, 2019)

In Africa, increased flooding and heavy rainfall often lead to the contamination of water supplies, fuelling outbreaks of cholera, typhoid fever, and other diarrheal diseases (Carlton et al., 2016; Kim et al., 2021). During the 2018 cholera outbreak in Zimbabwe, prolonged rainfall overwhelmed sanitation infrastructure, resulting in a rapid spread of *Vibrio cholerae* A( yling et al., 2023). Similarly, in Mozambique, Cyclone Idai (2019) led to massive flooding, triggering a large-scale cholera epidemic (Lequechane et al., 2020). The Durban floods of 2012 and 2022 had severe consequences for human health. The 2012 floods led to widespread water contamination, increasing cases of cholera, diarrhoea, and leptospirosis due to exposure to contaminated water sources(Ngcobo et al., 2023). The April 2022 floods, regarded as one of the deadliest climate disasters in South Africa’s history, resulted in over 435 fatalities and displaced thousands, exacerbating vulnerabilities in informal settlements (Olanrewaju & Reddy, 2023). The destruction of sanitation infrastructure and potable water supplies created ideal conditions for waterborne diseases, including typhoid fever and *E. coli*infections (Benschop et al., 2024). Additionally, stagnant water left in the aftermath facilitated mosquito breeding, heightening the risk of vector-borne diseases such as malaria and dengue fever (Meyer, 2024). The combined effects of displacement, overcrowding in emergency shelters, and inadequate access to healthcare services further intensified the spread of respiratory infections and other communicable diseases (Nyam et al., 2024). Conversely, drought conditions threaten water security, often forcing vulnerable populations to rely on unsafe drinking water sources, increasing the risk of gastrointestinal infections and waterborne illnesses (Kovats & Akhtar, 2008; Moyo et al., 2023) In East Africa, severe droughts have reduced access to potable water, leading to a spike in diarrheal diseases and acute malnutrition (Kitole et al., 2024). Prolonged droughts have been linked to increased concentrations of harmful algal blooms, contributing to toxic cyanobacteria outbreaks, which pose a threat to both human and animal health (Yuan et al., 2024). The link between environmental conditions and infectious disease outbreaks has garnered increasing attention in global health research, especially in regions that are both highly vulnerable to climate change and predisposed to public health crises(Haines et al., 2006; Liao et al., 2024; Semenza & Paz, 2021). South Africa faces a particular challenge with pathogen burden in waterways, which is often higher than expected due to inadequate water treatment and contamination from extreme weather events (Luyt et al., 2012; Potgieter et al., 2010).

The specific impact of EWEs on pathogen dynamics, particularly in relation to emerging and re-emerging infectious diseases, remains poorly understood (Grobusch & Grobusch, 2022). South Africa’s recent experiences with recurring droughts and floods, particularly in marginalized and rural communities, underscore the urgent need for systematic research to understand how climate change-driven EWEs influence infectious disease outbreaks (Hunter, 2003; Kapuka & Hlásny, 2021). Strengthening climate adaptation strategies, the need for environment-focused public health policies to mitigate the impact of climate change on disease dynamics and investing in public health infrastructure will be crucial in mitigating these risks and protecting vulnerable populations from climate-sensitive diseases.

## Aim

The aim of this systematic review is to understand emerging trends in disease outbreaks following extreme weather events in South Africa. This review addressed the research question registered on PROSPERO (Dickinson et al., 2024): ’What are the emerging trends in extreme weather events and infectious disease outbreaks in South Africa?’

Outcome measures:

1. What is the incidence of infection and what pathogens are causing outbreak(s)?
2. Who are the vulnerable and affected individuals, and do they have common characteristics (age, sex, underlying conditions, property ownership)?
3. What is the impact of EWEs such as flooding on management/communication strategies for emergent pathogen and contaminant and effective healthcare provider management?

This work provides the baseline to the project Warning system for Extreme weather events, Awareness Technology for Healthcare, Equitable delivery, and Resilience (WEATHER). - NIHR Funding and Awards (Lynch et al., 2025), informing the design of an early warning system for the potential of EWEs that will strengthen the resilience and action of affected communities and healthcare infrastructure and working practices, specifically in the Durban area of South Africa.

## Methods

The protocol for this systematic review was registered and published on PROSPERO following peer review (Dickinson et al., 2024).

*Search Strategy:* A comprehensive search strategy was developed using appropriate key words, free text terms and Boolean operators to maximize the retrieval of potentially relevant studies. The key words included: “South Africa” AND (“Extreme Weather Events” OR *alternate terms*) AND (“infectious disease” OR *alternate terms, including disease and pathogen nam*)*e*. *s*Full search terms can be found in Supplementary File 1. The search was conducted across various electronic databases for studies published between January 2014 – June 2024: Web of Science, PubMed, Science Direct, EBSCO (12 databases) and Cochrane Library.

*Eligibility Criteria:* Eligibility was based on the following inclusion and exclusion criteria:

## Inclusion criteria

The inclusion criteria encompass literature with a substantial focus on extreme weather events (EWEs) and infectious diseases, as well as research on the implementation of early warning systems. Eligible sources include peer-reviewed journal articles, government and non-governmental organization reports, and academic dissertations. A wide range of study designs is considered, including qualitative, quantitative, and mixed-methods studies, as well as systematic reviews, scoping reviews, meta-analyses, and rapid reviews.

## Exclusion criteria

The exclusion criteria include articles that were not published in English due to limited resources for translation. Additionally, studies that do not focus on extreme weather events (EWEs) and infectious diseases were excluded from consideration.

*Screening and Study Selection:* Screening and eligibility determination used a two-reviewer system (with consensus for disagreements and conferral with a third-party adjudicator if a consensus was unable to be reached). Studies were identified and screened by reviewing the title and abstract to remove all articles that clearly did not meet the eligibility criteria, followed by full text screening. Details about the included and excluded studies will be presented in a PRISMA diagram (Moher et al., 2009; Page et al., 2021).

*Data Extraction:* Four reviewers were involved in data extraction (ND, FH, LS, ML). The included articles were all reviewed and critically appraised independently by two reviewers, and data added to a data extraction table (Supplementary File 2).

*Methodological quality assessment:* All studies were rated for quality using study-specific quality appraisal tools from the Joanna Briggs institute (Moola et al., 2017; Munn et al., 2021). The authors adopted a scoring system to rate quality as high, medium or low.

*Data Synthesis:* An overarching narrative synthesis was conducted across the evidence to form a structured narrative of results. Narrative synthesis was carried out following the guidelines of the Cochrane Collaborative (https://cccrg.cochrane.org/sites/cccrg.cochrane.org/files/uploads/AnalysisRestyled.pdf). Themes emerged inductively from examination of the data extraction table and ongoing discussions between the authors.

## Results

The PRISMA diagram (Figure 1) visually represents the process of identifying, screening and including studies in this review (Page et al., 2021).

**Fig. 1.**
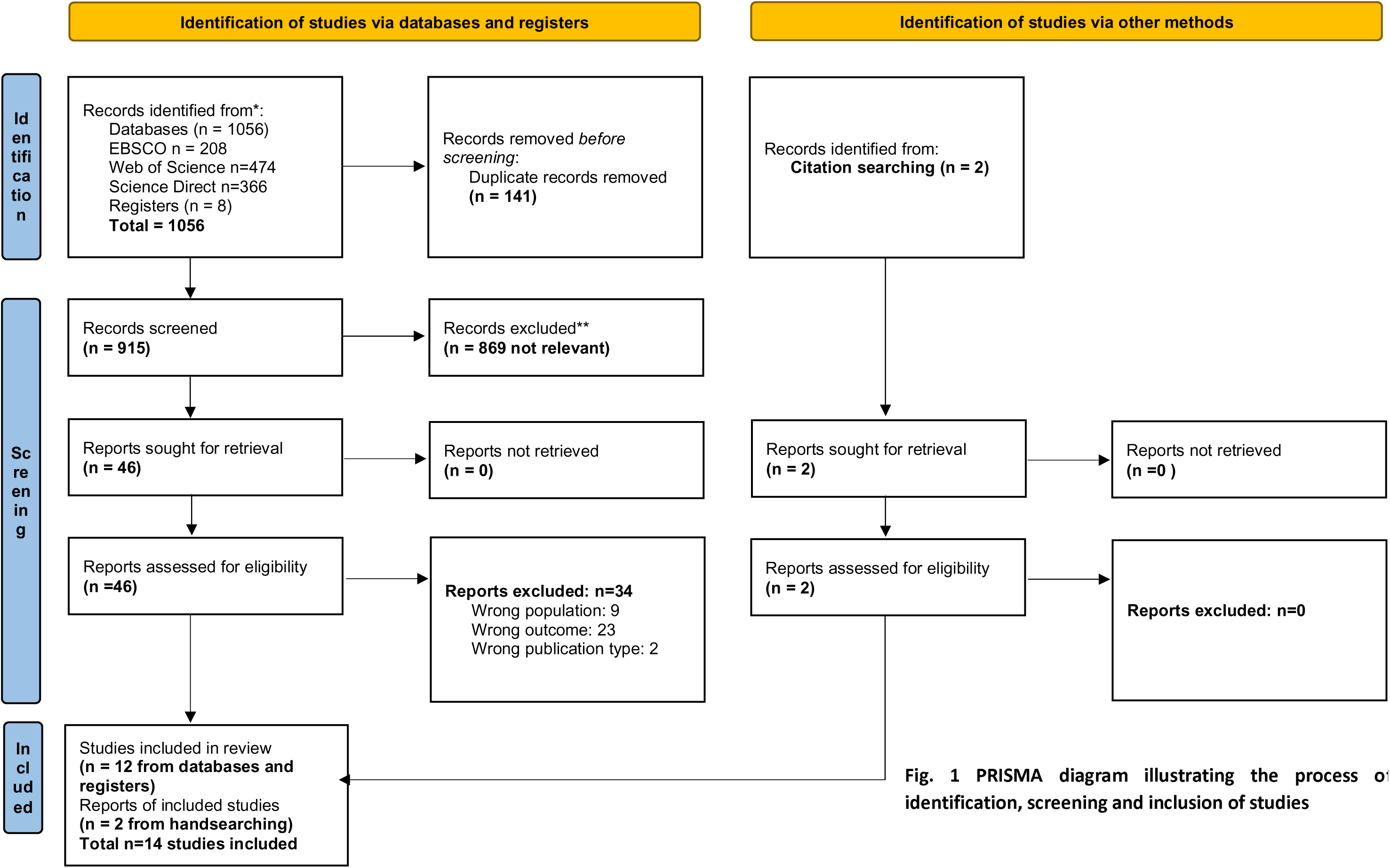
PRISMA diagram illustrating the process o identification, screening and inclusion of studies

Fourteen studies were included in the systematic review and defined in types of study (Figure 2). Table 1 presents summary data of the included studies. The full data extraction and critical appraisal information can be found in Supplementary File 2.

**Fig. 2.**
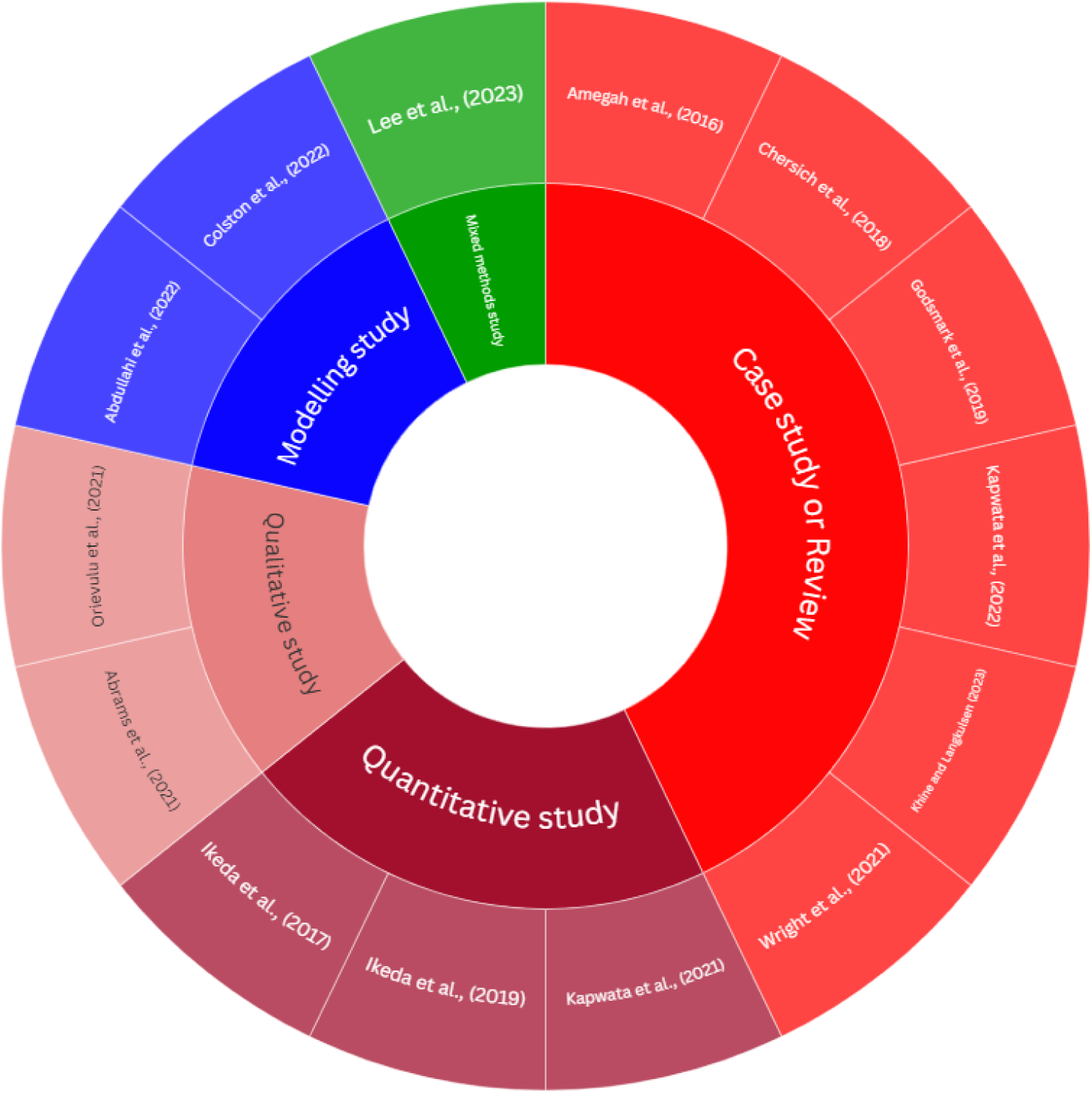
Evidence map of the included studies by study type

**Table 1:**
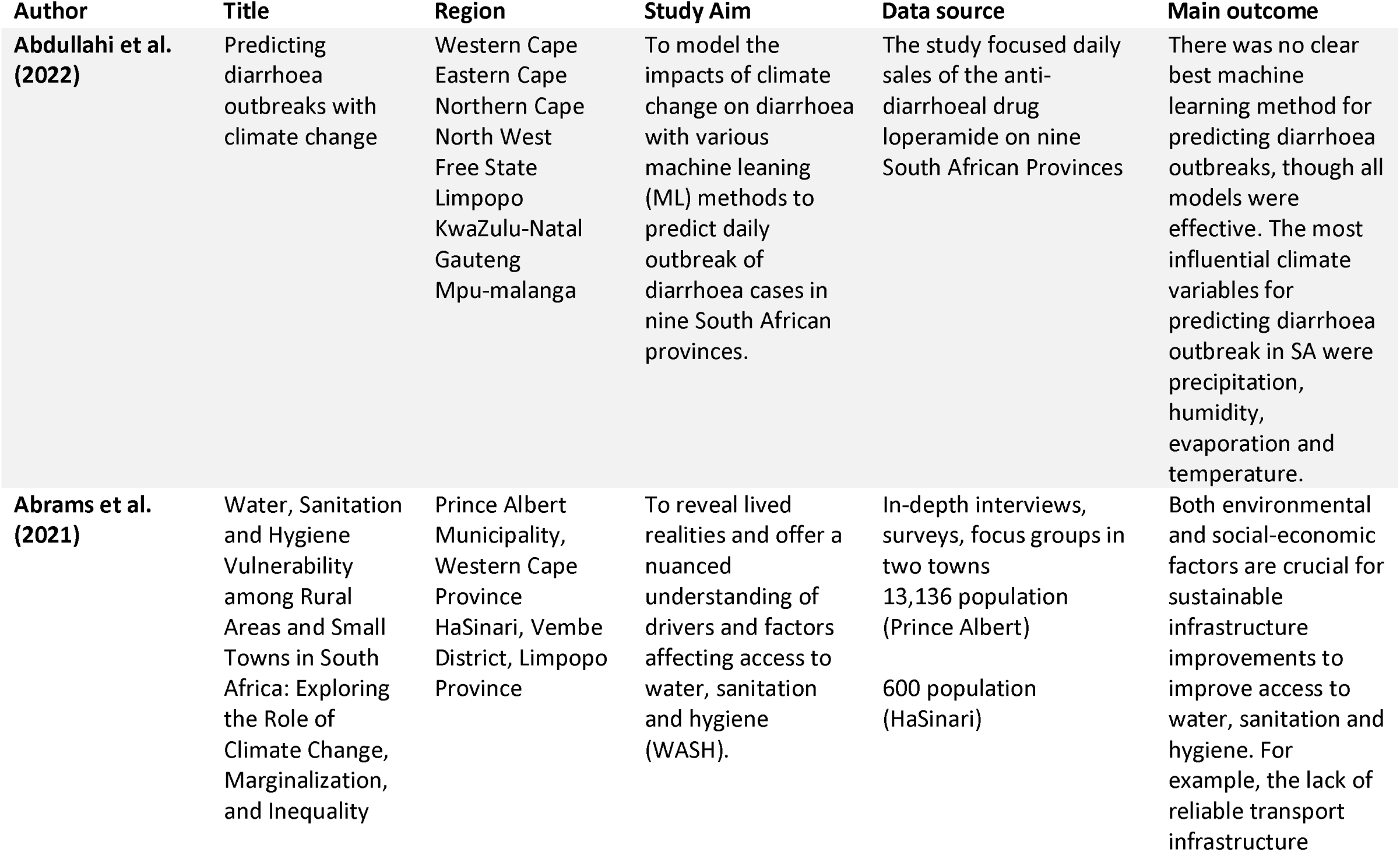
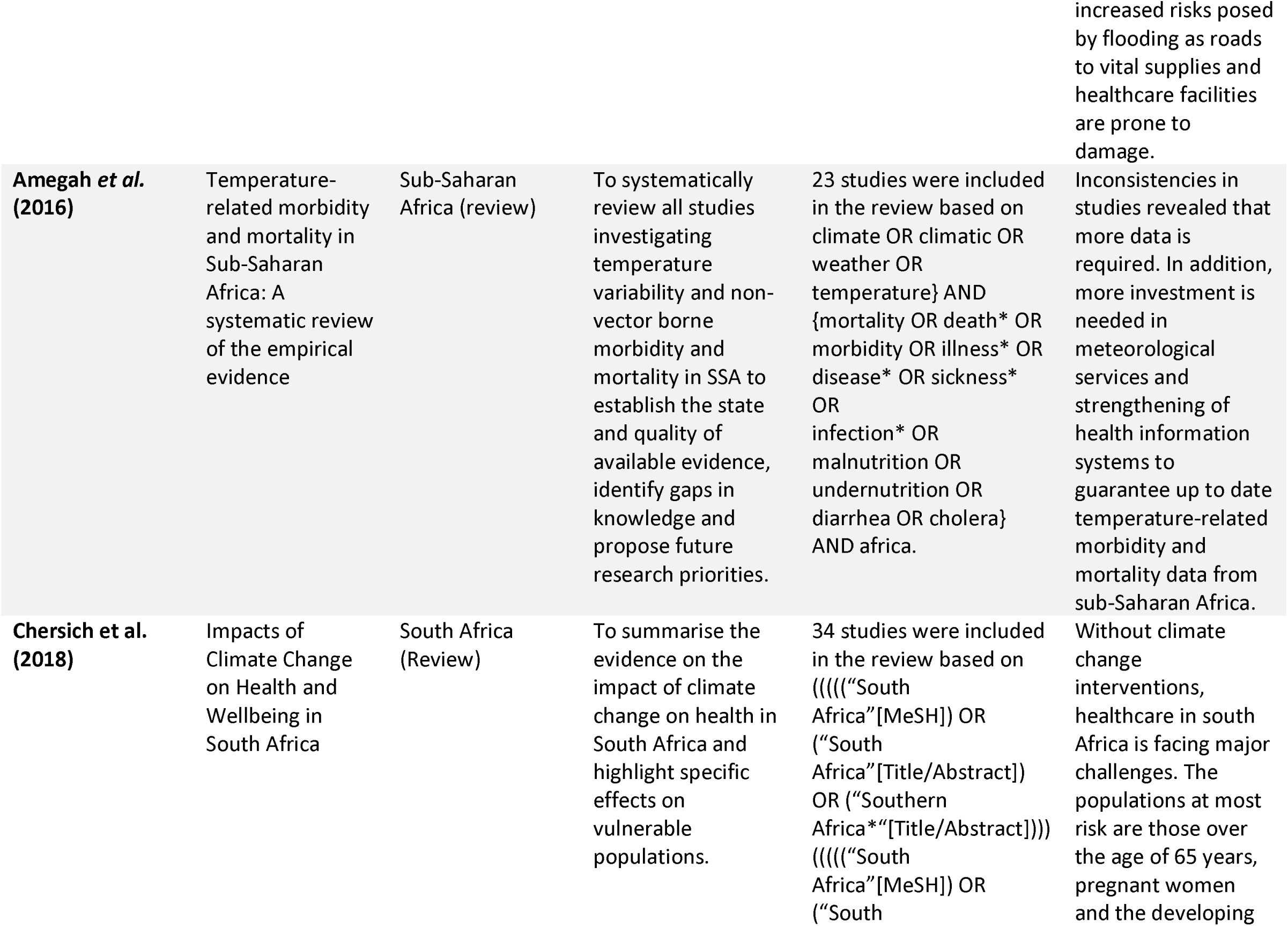
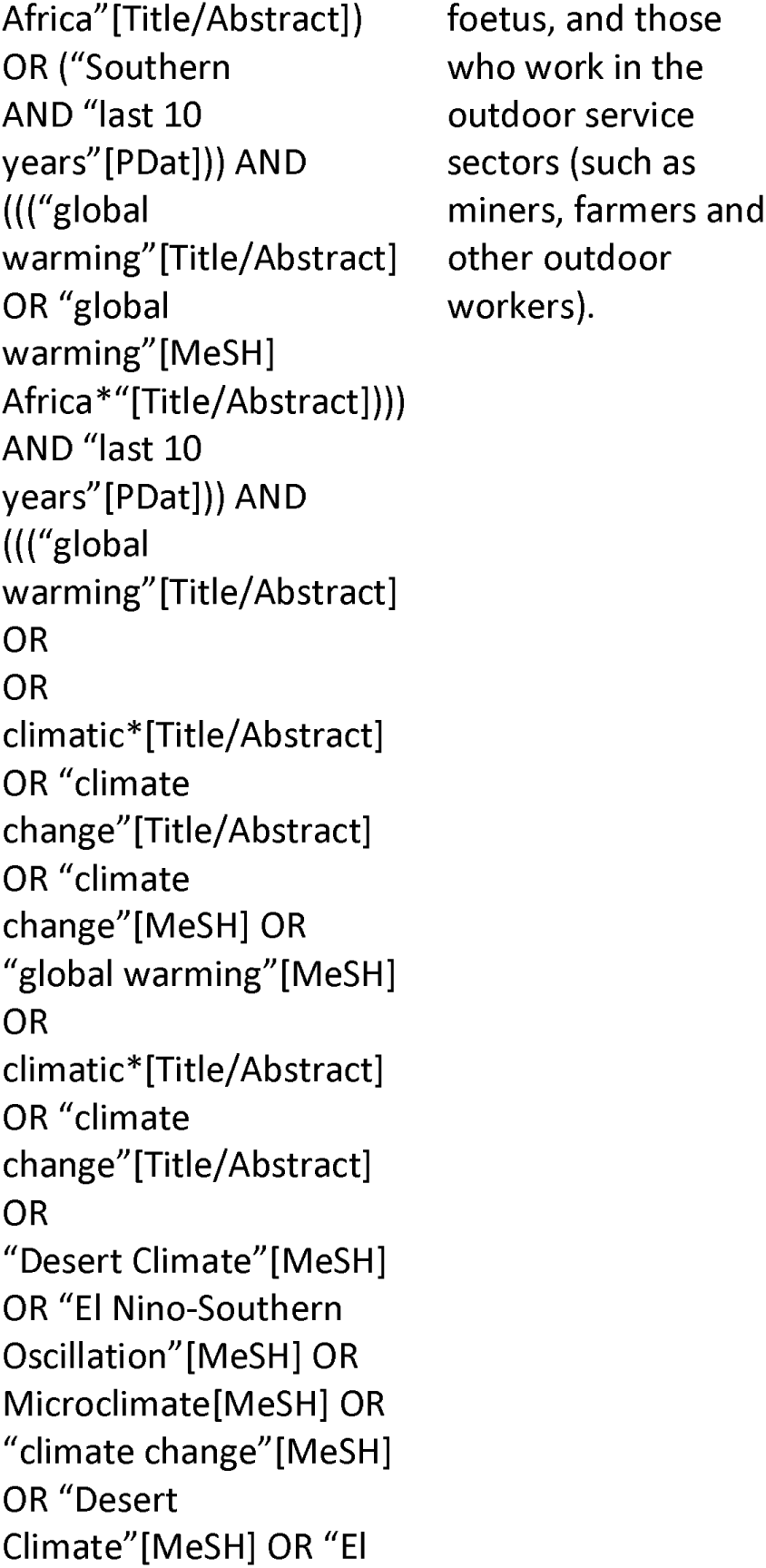
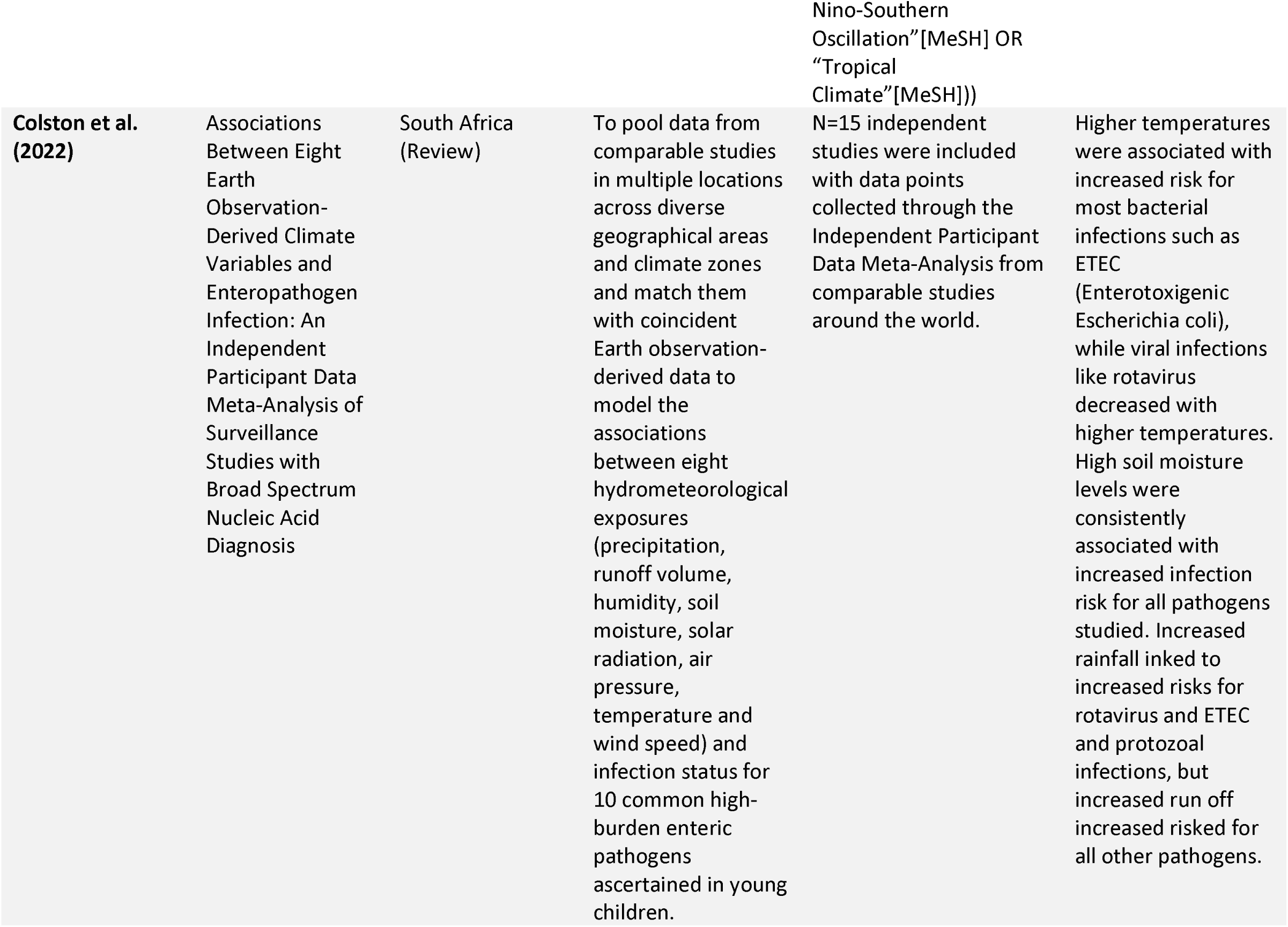
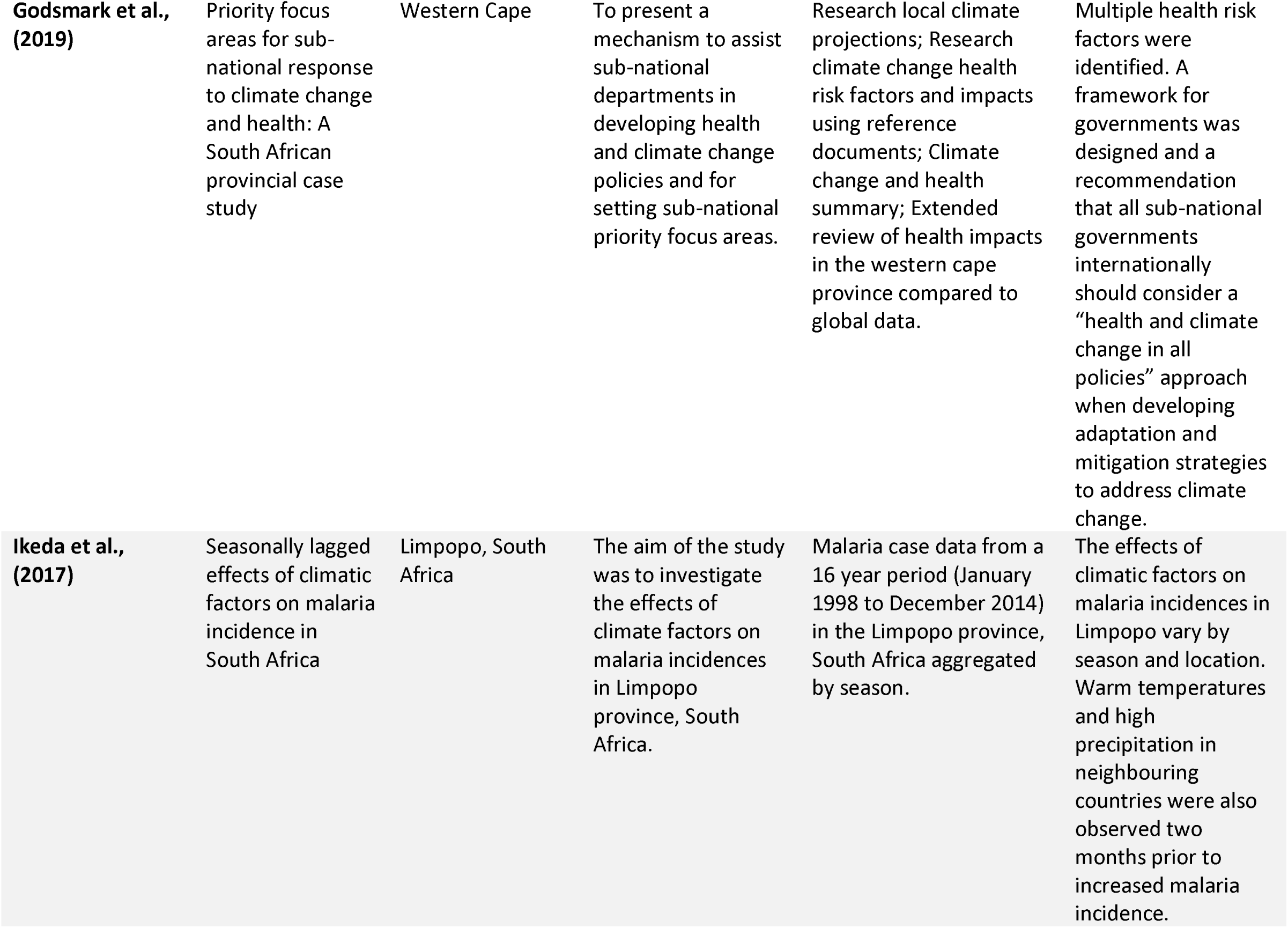
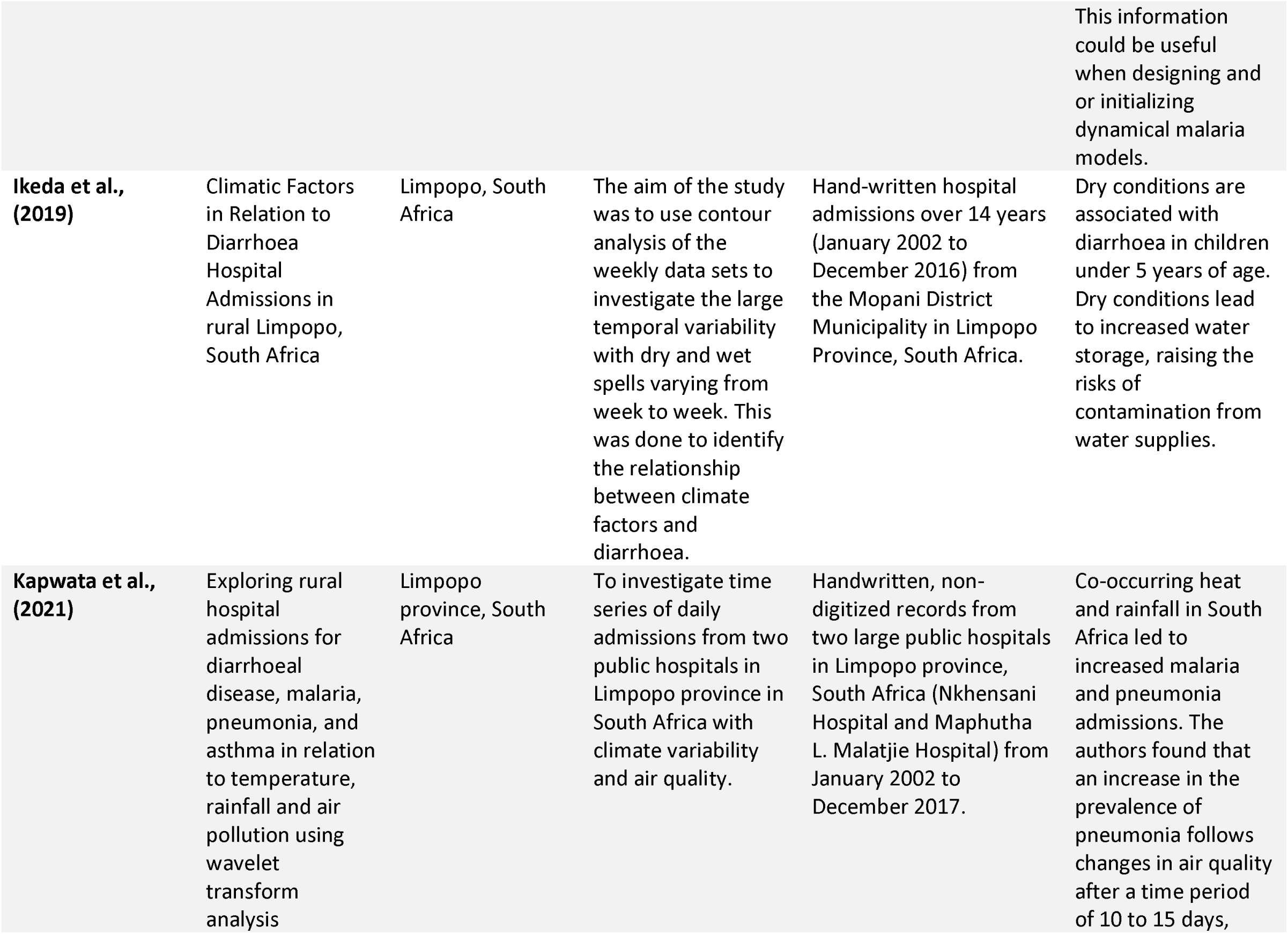
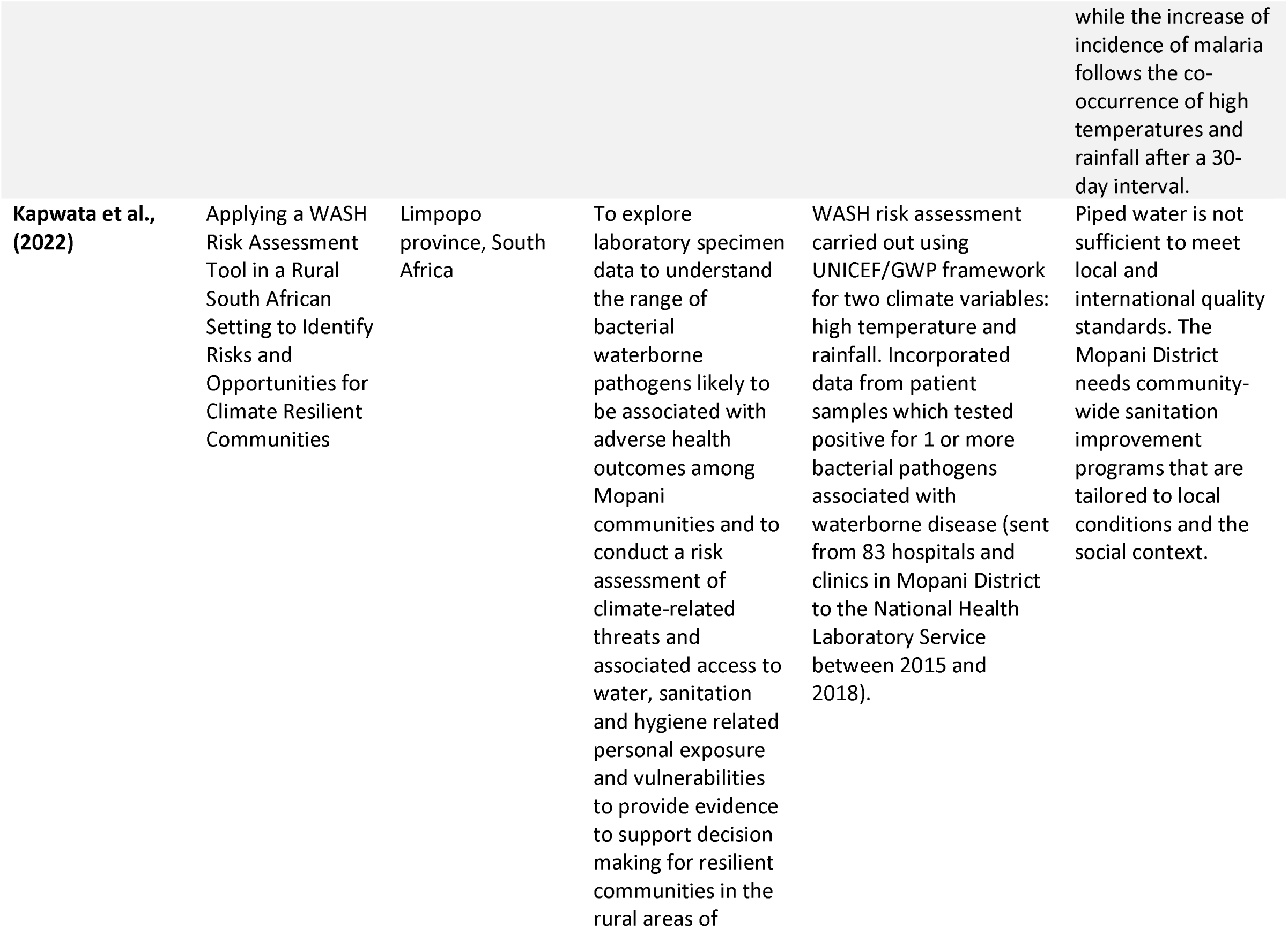
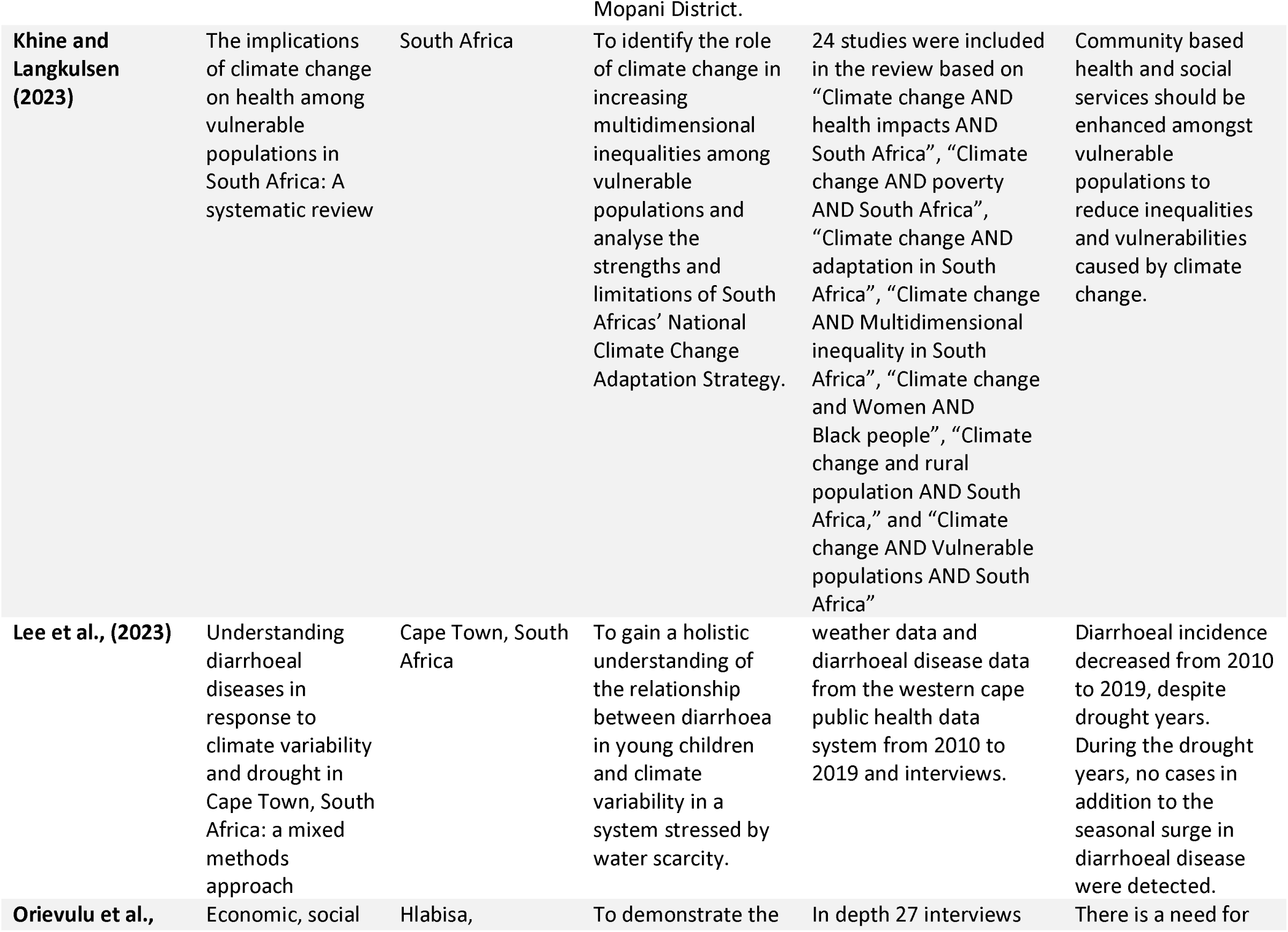
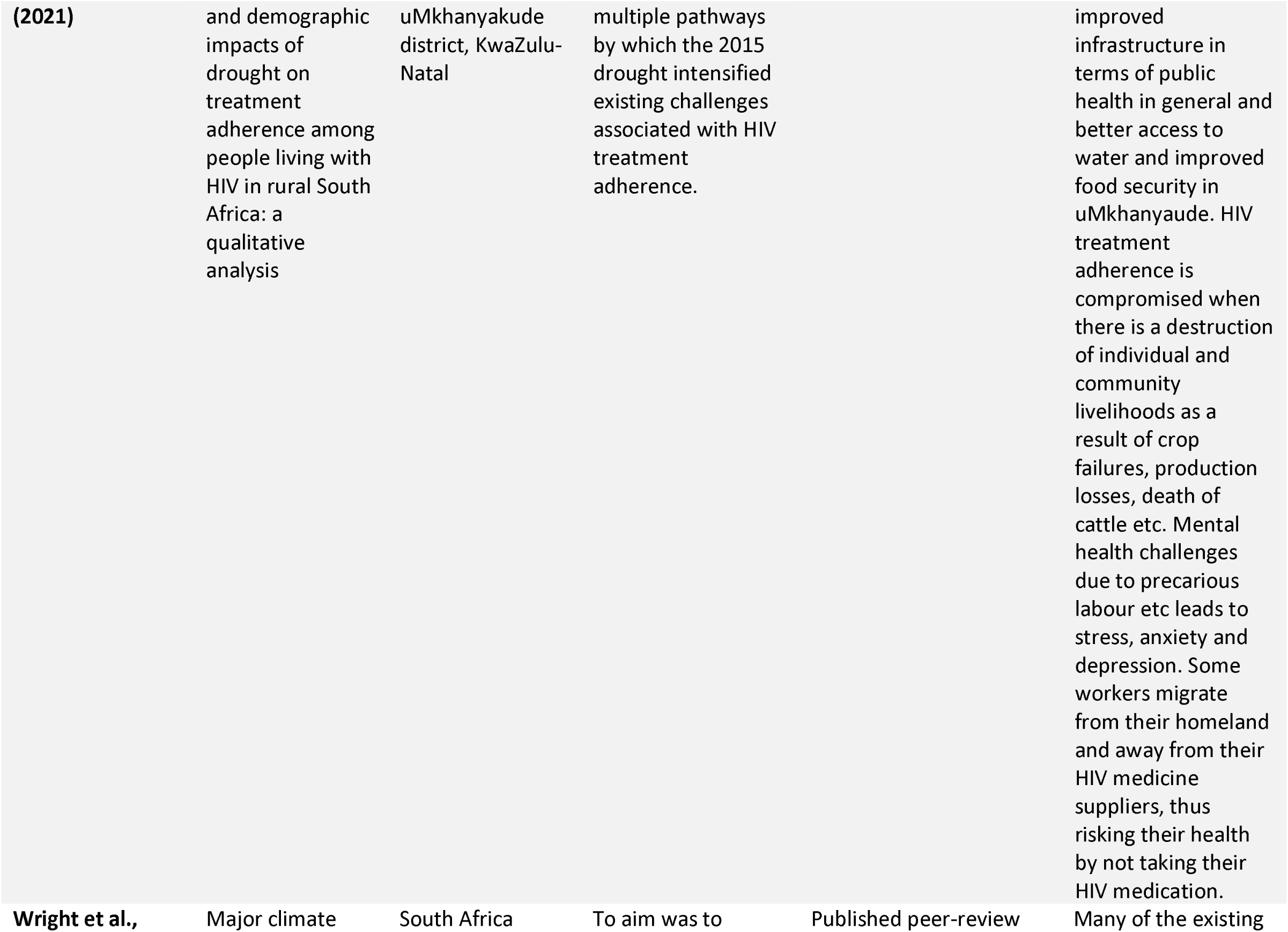
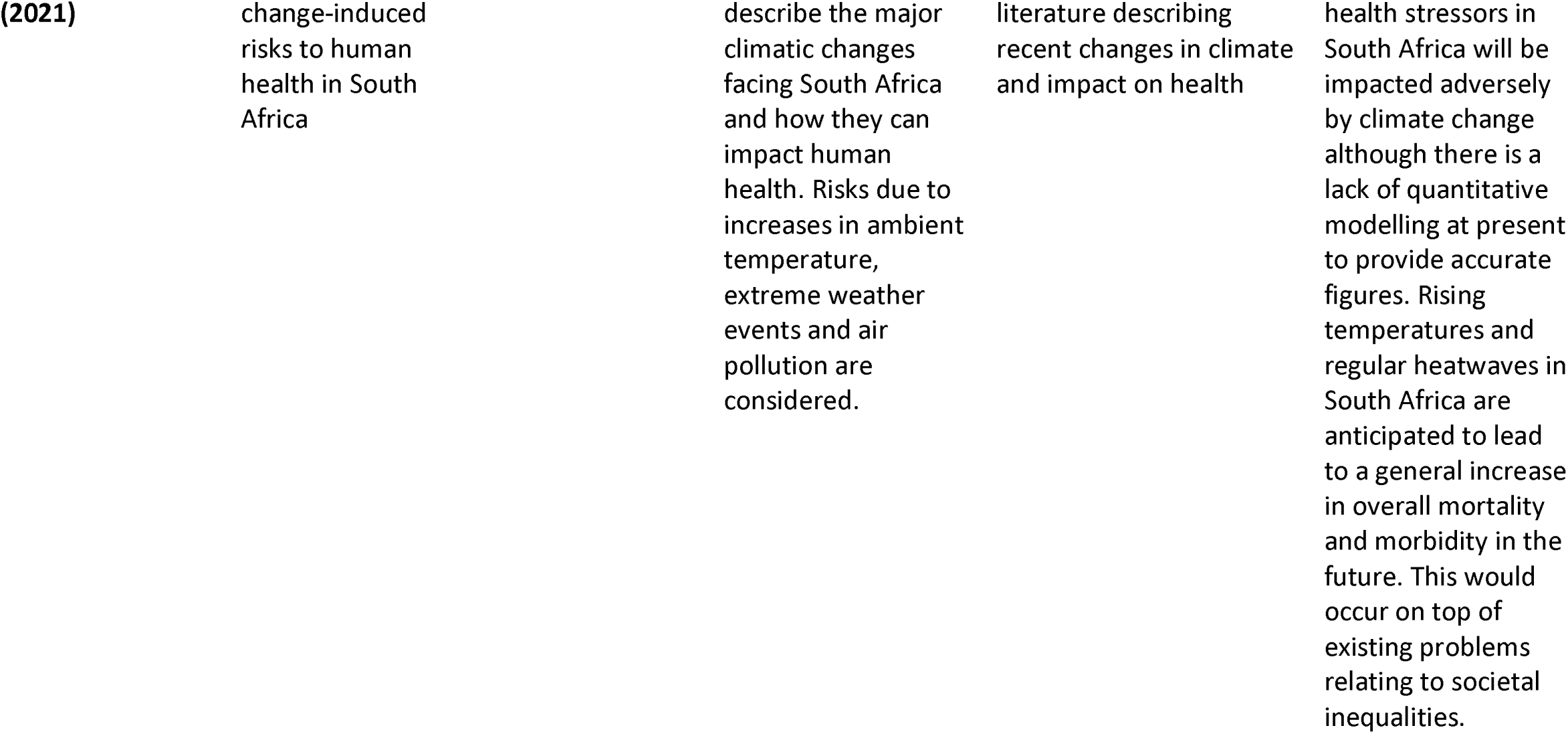
Study characteristics.

Despite the paucity of diagnostic datasets and environmental monitoring it is possible to identify an increase in infectious disease after an EWE and no difference of EWE type (Table 2).

**Table 2:** Summary of impact of weather variables on causative pathogens to infectious disease.

| Category | Pathogens | Symptoms | Favourable Weather Variables |
| --- | --- | --- | --- |
| **Environmental**<br>**Gastro-intestinal** | Bacteria<br>*Escherichia coli*<br>*Vibrio cholera*<br>*Salmonella* species<br>Virus<br>Rotavirus<br>Protozoa<br>*Cryptosporidium*<br>*Giardia* | Diarrhoea | ↑Temperature<br>↑Humidity<br>↑Rainfall<br>↑Soil moisture<br>↓Temperature<br>↑Soil moisture<br>↑Temperature<br>↑Soil moisture |
| **Environmental**<br>**Respiratory** | Bacteria<br>*Mycobacterium tuberculosis*<br>Virus<br>Respiratory viruses | Cough<br>Chest infection<br>Fever | ↓Temperature<br>↑Rainfall (Flooding leading to overcrowding and population proximity) |
| **Vector-borne**<br>**Mosquito** | Virus<br>Dengue<br>Protozoa<br>*Plasmodium* (malaria) | Fever<br>Malaise | ↑Temperature<br>↑Rainfall |
| **Vector-borne**<br>**Snail** | Schistosomiasis | Urogenital<br>GI<br>Liver | ↑Humidity<br>↑ Rainfall |

Key: ↑ = increase; ↓ = decrease

Through a critical analysis of studies included three themes emerged inductively addressing the influences of the emergence of infectious disease outbreaks following EWEs in SA.

## Themes

1. ‘Climate impacts on infectious disease’: climatic impacts on infectious disease and causative pathogens highlight how shifts in temperature, humidity, and precipitation influence disease transmission, particularly through waterborne and vector-borne pathogens.
2. ‘Population vulnerability’: marginalized communities, including those in informal settlements, face disproportionate health risks due to overcrowding, poor sanitation, and limited healthcare access.
3. ‘Policy and infrastructure’: - the destruction of water treatment plants, sewage systems, and healthcare facilities compounds public health crises.

## Discussion

### Theme 1: Climate impacts on infectious disease

While most climate and health research in South Africa has focused on long-term climate change trends, there is growing interest in the short-term impacts of EWEs. The evidence highlights that drought (exceptionally low rainfall) and flooding (exceptionally high rainfall) are the most significant climatic factors associated with infectious disease outbreaks in South Africa. Studies have established links between rising temperatures, EWE’s such as flooding, heavy rainfall, drought, the El Niño Southern Oscillation, and hurricanes, and the incidence of waterborne and enteric infections including hepatitis, rotavirus, norovirus, enterovirus, cholera, giardia, typhoid, and Legionnaires’ disease (Godsmark et al., 2019).

### Waterborne Diseases and Sanitation Challenges

Diarrheal diseases are among the most frequently discussed infections in relation to EWEs (Abdullahi et al., 2022; Ikeda et al., 2019; Lee et al., 2023). These are highly sensitive to rising temperatures, and outbreaks often **i**ntensify during droughts and flooding due to poor sanitation, hygiene challenges, and deteriorating water quality (Abrams et al., 2021). A longitudinal study of diarrhoeal admissions and weather variables found lagged admissions for diarrhoeal disease were associated with wetter than average conditions, but not significantly related to temperature in those over 5 years of age, whilst in under 5’s, diarrhoeal admissions were associated with both drier than average and wetter than average conditions, as well as high temperatures (Ikeda et al., 2019). Behavioural changes are critical in reducing disease transmission during extreme weather events. For instance, water storage practices during drought conditions significantly impact the risk of diarrheal diseases, increased water storage, often necessary due to water scarcity, can inadvertently lead to higher contamination risks, contributing to increased diarrhoea rates (Ikeda et al., 2019). Additionally, reduced water availability leads to poor personal hygiene, exacerbating the spread of diseases which thrive in poor sanitation environments (Abrams et al., 2021). A mixed-methods study by Lee et al. (2023) in Cape Town found a 64.7% reduction in diarrheal cases with dehydration over a 10-year period (2010–2019) despite drought years. During the drought years, no significant increase in cases over and above the seasonal surge in diarrhoeal disease were detected. Evidence suggests that public health interventions, including rotavirus immunization and consistent Water, Sanitation, and Hygiene (WaSH) messaging, contributed to this decline (Lee et al., 2023). However, despite this progress, drought conditions remain a major risk factor for diarrheal disease due to water scarcity, unsafe water storage, and reduced personal hygiene practices (Ikeda et al., 2017; Lee et al., 2023). This underscores the need for targeted interventions to enhance WaSH infrastructure and practices during extreme weather events.

### Vector-Borne Diseases

Vector-borne infections are particularly sensitive to long-term climate trends, with projections indicating a significant increase in mosquito-borne infections in the future(Wright et al., 2021). Rising temperatures and increased rainfall enhance vector survival, prolong transmission seasons, and expand the geographical range of these diseases. For instance, schistosomiasis and mosquito-borne infections are expected to move into previously unsuitable environments as climate patterns shift toward warmer and wetter conditions, creating optimal conditions for vector proliferation (Wright et al., 2021). A cohort study of seasonally lagged effects of climate variables on malaria incidence observed associations between both cool air temperatures over southern Africa (La Niña) and high precipitation and high temperatures in neighbouring countries, suggesting the need to consider not only local but also more distant climatic conditions (Ikeda et al., 2017).

However, Amegah et al. (2016) critique time-lag studies, particularly those using longer timescales and time lags of up to 6 months (Ikeda et al., 2017)as it is difficult to differentiate between seasonal patterns and short-term variation, and recommend time series regression as the preferred method to investigate climate and health relationships (Amegah et al., 2016).

### Infectious Disease Seasonality and Extreme Weather Events

Schistosomiasis, malaria, and other mosquito- or tick-borne diseases, exhibit annual or biannual seasonal trends, with peak transmission occurring in wet, summer months (Wright et al., 2021). However, EWEs can also disrupt typical seasonal patterns, leading to unexpected disease spikes. For example, malaria and enterotoxigenic *E. coli* (ETEC) infections have been shown to surge following extremes in rainfall and temperature (Colston et al., 2022). Among EWE-associated infectious diseases in SA, malaria and diarrheal diseases are the most frequently studied, with pneumonia, linked to air-borne viruses emerging as a significant concern. Studies indicate that co-occurring high temperatures and precipitation are associated with increases in malaria and pneumonia cases (Ikeda et al., 2017; Kapwata et al., 2021).

### Machine Learning and Disease Prediction

Predicting diarrheal disease outbreaks has been a focus of recent research. Abdullahi et al. (2022) found that Convolutional Neural Networks (CNNs), a deep learning method, were the most statistically effective predictive models (Abdullahi et al., 2022). However, it is worth noting that diarrhoea cases over a ten-year period were estimated using sales of Loperimide as a proxy, as opposed to healthcare records. Key climate variables — precipitation, humidity, evaporation, and temperature — were identified as the most influential in forecasting diarrheal outbreaks. These findings support the development of early warning systems for climate-sensitive diseases, demonstrating the potential of machine learning (ML) models in public health planning. Similarly, a meta-analysis, pooling data from multiple studies, including two from SA and six from sub-Saharan Africa (Colston et al., 2022) matched climate data with infectious pathogen outbreaks, revealing that bacterial infections tend to increase with higher humidity, while viral infections decline in warmer temperatures. Soil moisture was identified as a key risk factor for all enteropathogens, though the impact of precipitation was mixed. Future research should focus on pathogen-specific seasonal trends at a localized scale to refine disease forecasting models (Colston et al., 2022)

In summary, the same types of infectious diseases are found in flood and drought in SA. The pathogens that influence disease outbreaks are mobilised or concentrated by EWEs by providing improved conditions for pathogen transmission and proliferation in the environment. In agreement with Amegah et al.’s (2016) recommendation, future work should focus on the characterisation of the outbreaks, linking cases of diseases to thorough testing in the population to identify asymptomatic carriers of specific pathogens (Amegah et al., 2016).This would enable a reliable dataset for pathogen toolbox to be utilised as part of a robust approach to predict EWEs, significant changes in rainfall patterns and general climate change preparedness.

### Theme 2: Population vulnerability (dealing with EWEs)

The health impacts of EWEs are linked to human behavioural and socio-economic factors, with those who are more vulnerable more likely to experience disease (Khine & Langkulsen, 2023). The occurrence of EWEs intensifies the existing challenges experienced by the population (Orievulu et al., 2022). Vulnerable groups have been highlighted as a priority area for sub-national government focus in relation to EWEs (Godsmark et al., 2019). Due to the high level of underlying morbidity, financial insecurity, poor-quality housing and sanitation, and unreliable access to healthcare, the level of vulnerability in the South African population is high (Wright et al., 2021).

The populations most at risk in SA are those over the age of 65 years, pregnant women and the developing foetus, children under 5 years of age, those who work in the outdoor service sectors (such as miners, farmers and other outdoor workers), those with pre-existing medical conditions, those living in poverty, displaced populations and those living in informal settlements (Chersich et al., 2018; Godsmark et al., 2019; Wright et al., 2021). Khine and Langkulsen (2023) also highlight that women who have a higher poverty rate are more susceptible to climate change induced health effects, particularly climate-related disasters; the indirect health effects of EWEs is compounded in women due to systemic discrimination, unequal access to resources, and unequal roles and responsibilities than males (Khine & Langkulsen, 2023). In addition, Godsmark et al., (2019) draw attention to the vulnerability of women and children in displaced communities, where lack of privacy in makeshift shelters and a breakdown of social structures have been found to result in increased sexual abuse (Godsmark et al., 2019). To reduce vulnerabilities and enhance resilience, authors suggest expanding community-based health and social services, particularly in rural and marginalized communities, which are disproportionately affected by EWEs (Khine & Langkulsen, 2023).

Whilst EWEs do not directly affect the HIV virus, vulnerabilities in the population living with the HIV virus are exacerbated due to EWEs. Evidence examining a systems approach identified a complex, multidimensional pathway through which drought impacts on HIV treatment adherence. Results indicate that drought-induced water shortages (causing reduced access to water and food, loss of livestock and reduced agricultural production) led to disruptions to livelihoods, income, food systems, and general health. The additional stress placed on vulnerable populations was determined to impact on HIV treatment adherence, through a combination of compromised access to medicines and healthcare, lack of water with which to take pills, and stress-induced reduction in health-seeking behaviours (Orievulu et al., 2022). In addition, during periods of heat stress, access to healthcare and medication is impacted, leading to non-adherence to prescribed treatment and an associated rise in viral load. In breastfeeding women, the risk of mother-to-child transmission increases as hot weather leads to higher intake of HIV-containing breast milk by the infant. (Wright et al., 2021).

Furthermore, the impact of social displacement and disruption, alongside gender disparities, in undermining both the prevention and treatment of HIV (Chersich et al., 2018). Local living conditions and environmental health factors are likely to overshadow typical climate-related patterns influencing diarrheal diseases (Ikeda et al., 2019). There is increased transmission of infectious disease in environmental refugees, due to overcrowding, poor sanitation and inadequate access to clean drinking water (Godsmark et al., 2019). Continuous access to clean water is a critical factor in reducing vulnerability to infectious diseases. Examination of two distinct regions of SA to barriers and vulnerabilities with regards to water, sanitation and hygiene (WaSH), identified that the WaSH services in rural and small towns being influenced by a range of factors, including history (inequalities remain from the Apartheid era), natural environment, socio-economic and infrastructure challenges (Abrams et al., 2021).

### Theme 3: Policy and infrastructure: Gaps, Successes and Future Tools

EWE’s in South Africa are expected to become more frequent and severe, necessitating a stronger integration of health considerations into climate adaptation and mitigation strategies that encompass infectious disease in addition to other multidimensional risk factors (Chersich et al., 2018). Policy on climate change and health is lacking at sub-national level, and suggest that provincial governments should adopt “health and climate change in all policies” approach to ensure that climate resilience is embedded across multiple sectors, including health, infrastructure, and disaster response (Godsmark et al., 2019).

Research conducted in the Western Cape identified several under-addressed critical health impacts, including water-borne and food-borne diseases, and mental health (Godsmark et al., 2019). However, despite the potential severity of these health impacts, historically health impacts have received limited attention in South Africa’s sub-national climate planning. Nevertheless, the South Africa’s National Climate Change Adaptation Strategy (Department of Environment Forestry and Fisheries Republic of South Africa, 2019)includes some health-focused measures, there is a lack of emphasis on mental and occupational health, which are critical components of climate adaptation) (Khine & Langkulsen, 2023). This omission of focus on mental health is significant because conditions such as stress, anxiety, and post-traumatic stress disorder are common following extreme weather disasters. Evidence identified that occupational health risks associated with rising temperatures and environmental hazards (e.g., agricultural workers’ exposure to heat stress and pesticide poisoning) require greater policy attention (Khine & Langkulsen, 2023). Addressing these gaps will require a holistic, multi-sectoral approach that integrates public health, social services, and infrastructure planning (Kapwata et al., 2022).

The impacts that flooding can impact on accessing healthcare facilities following EWEs, due to lack of reliable transport infrastructure in combination with damage to and flooding of roads, resulting in loss of or reduced access to vital supplies, including medicines. Inadequate transport infrastructure can also result in flooded populations being unable to access healthcare facilities. In addition, Water, Sanitation and Hygiene vulnerability (WaSH) in South Africa are affected by community practices, hygiene practices and cultural beliefs (Abrams et al., 2021). The use of water sources for multiple purposes (e.g. washing, drinking and for livestock) can affect water availability and quality. Cultural beliefs and education may influence how communities manage and prioritize WaSH resources and how they respond to WaSH challenges(Abrams et al., 2021). In addition, the evidence suggests that while improving sanitation infrastructure and ensuring access to clean water are key interventions, behavioural factors — such as illegal water connections, poor storage practices, and inconsistent hygiene habits — also play a role in maintaining water safety. The results further highlights that the presence of WaSH facilities does not necessarily mean the infrastructure will be functional. Upgrading water distribution systems (e.g., enhancing pipe quality) and cracking down on unauthorized water connections can improve water quality and availability (Ikeda et al., 2019).

The health service in SA have now focused on maintaining databases to allow accurate tracking of climate-sensitive diseases, as was called for as far back as 2004 in the National Climate Change Response Strategy(Department of Environmental Affairs and Tourism, 2004)called for the development of early warning systems for infectious diseases, heat stress and air pollution exposure. This call was strengthened in the National Climate Change Adaptation Strategy (Department of Environment Forestry and Fisheries Republic of South Africa, 2019). There have been challenges in the healthcare system which have delayed this, including the often-lacking data from primary healthcare facilities (Wright et al., 2021). Additionally, Abdullahi et al., (2022) recognise that patients presenting to healthcare facilities with diarrhoea tend to be treated for symptoms, but stool samples are not routinely tested to determine the pathogens causing the infection; this means limited data is available to allow modelling of specific pathogen burden relating to EWEs (Abdullahi et al., 2022). There is evidence of further support this view and a realisation that that very few studies have used empirical health service data to analyse climate-related impacts on health (Chersich et al., 2018).These results are not in isolation with Amegah et al.’s (2016) study examining temperature related mortality and morbidity found inconsistencies in the evidence suggesting a need for additional data, as well as investment in meteorological and health services (Amegah et al., 2016).

The emerging evidence suggests there is a need to reframe climate change as a health issue in SA, and that healthcare professionals have the potential to be key players in identifying and recording the impacts of climate change on health, and supporting mitigation strategies (Chersich et al., 2018). There have been recommendations that content on climate change and risks to health (including measures individuals can take to protect themselves) be included in the National Curriculum for education, and that higher education programmes for teachers and healthcare professionals should incorporate specific training on this (Wright et al., 2021).

## Conclusions

This review investigated emerging trends between extreme weather events (EWEs) and infectious diseases, identifying an increase in disease outbreaks regardless of the type of EWE (flood or drought). Three key themes emerged from the analysis. First, climatic impacts on infectious diseases highlight how temperature, humidity, and precipitation influence transmission, particularly of waterborne and vector-borne pathogens. Increased rainfall and flooding often contaminate water sources, facilitating outbreaks of diseases such as cholera and leptospirosis, while stagnant water promotes mosquito breeding, exacerbating malaria and dengue fever risks.

Second, population vulnerabilities emphasize that marginalized communities, particularly those in informal settlements, face disproportionate health risks due to overcrowding, poor sanitation, and limited healthcare access. Displacement following EWEs increases exposure to communicable infectious diseases, particularly respiratory and gastrointestinal infections. Vulnerable groups identified in South Africa mirror those from a recent UK systematic review (Dickinson, Spencer, et al., 2024), underscoring that socio-economic and health disparities drive climate-related health impacts worldwide.

Finally, infrastructure plays a critical role in post-EWE recovery, as the destruction of water treatment plants, sewage systems, and healthcare facilities compounds public health crises. The review underscores the need for climate-resilient health systems, improved emergency preparedness, and targeted interventions to mitigate the health burden of EWEs.

There is a lack of data on pathogen environmental distribution following EWEs. While South Africa faces significant health challenges due to increasing EWEs, strategic planning, infrastructure investment, and community engagement can help mitigate risks. Implementing AI-driven predictive tools, such as climate-health early warning systems, offers an opportunity to protect communities and prepare health services. A machine-learning study in South Africa suggests that additional environmental variables, such as sea surface temperature, may enhance malaria prediction over land-based weather measures (Martineau et al., 2022). Similarly, precipitation in neighboring countries influences lagged malaria incidence (Ikeda et al., 2017), highlighting the need to consider broader variables in outbreak prediction.

Addressing socio-ecological factors, improving disease surveillance, and integrating climate-health training will be essential for building climate-resilient healthcare systems. Effective data collection and framing climate change as a health issue require increased leadership from the health sector (Chersich et al., 2018). Health system development must incorporate planetary health into policy and practice, with curriculum enhancements recommended for healthcare professionals (Irlam et al., 2024). Community-based health services should address multidimensional vulnerabilities, while monitoring pathogen exposure during flood clean-up efforts remains crucial (Dickinson, Spencer, et al., 2024).

This review aligns with previous research (Destoumieux-Garzón et al., 2022) emphasizing the importance of a One Health approach to investigating climate-driven infectious diseases. Bridging policy gaps and leveraging technology will enable South Africa to develop proactive public health strategies, including improved water infrastructure and early warning systems, to mitigate the impact of climate-sensitive infectious diseases (Ayling et al., 2023).

## Recommendations for Research, Policy and Practice

From the available data the recommendations for research, policy and practice.

- Large longitudinal datasets are needed to understand the dynamics of specific pathogens, particularly waterborne pathogens, during and after Early Warning Systems in SA, to inform predictive models.
- Public health organisations should prioritise pathogen diagnostics in diarrhoea hospital admissions and seek information regarding flooding status of their home/community areas.
- The education sector and healthcare sector should work collaboratively to ensure both the general public and healthcare professionals are educated on the interrelation of EWEs and health, and subsequent strategies to mitigate disease risk.
- Early Warning Systems should incorporate data on precipitation, humidity, evaporation and temperature as these variables are most associated with increased disease risk.
- An Early Warning System should be developed to warn communities about EWE’s such as flooding, enabling individuals and families to stay safe and enabling removal of property and livestock from flood level areas.
- Local health services should be warned about extreme weather events for them to effectively plan delivery of usual care and emergency health and social care in the aftermath of a flood or drought.

## Data Availability

All data produced in the present study are available upon reasonable request to the authors

## References

1. Abdullahi, T., Nitschke, G., & Sweijd, N. (2022). Predicting diarrhoea outbreaks with climate change. PLoS ONE, 17(4 April). 10.1371/journal.pone.0262008

2. Abrams, A. L., Carden, K., Teta, C., & Wågsæther, K. (2021). Water, sanitation, and hygiene vulnerability among rural areas and small towns in south africa: Exploring the role of climate change, marginalization, and inequality. Water (Switzerland), 13(20). 10.3390/w13202810

3. Acosta-España, J. D., Romero-Alvarez, D., Luna, C., & Rodriguez-Morales, A. J. (2024). Infectious disease outbreaks in the wake of natural flood disasters: global patterns and local implications. In Infezioni in Medicina(Vol. 32, Issue 4, pp. 451–462). EDIMES Edizioni Medico Scientifiche. 10.53854/liim-3204-4

4. Amegah, A. K., Rezza, G., & Jaakkola, J. J. K. (2016). Temperature-related morbidity and mortality in Sub-Saharan Africa: A systematic review of the empirical evidence. In Environment International (Vol. 91, pp. 133–149). Elsevier Ltd. 10.1016/j.envint.2016.02.027

5. Ayling, S., Milusheva, S., Kashangura, F. M., Hoo, Y. R., Sturrock, H., & Joseph, G. (2023). A stitch in time: The importance of water and sanitation services (WSS) infrastructure maintenance for cholera risk. A geospatial analysis in Harare, Zimbabwe. *PLoS Neglected Tropical Disease*,s17(6 June). 10.1371/journal.pntd.0011353

6. Benschop, N., Chironda-Chikanya, G., Naidoo, S., Jafta, N., Ramsay, L., & Naidoo, R. (2024). El Niño, Rainfall and Temperature Patterns Influence Perinatal Mortality in South Africa: Health Services Preparedness and Resilience in a Changing Clima(tpep. 333–355). https://link.springer.com/chapter/10.1007/978-3-031-38878-1_21#:~:text=Our%20data%20provides%20evidence%20that,West%20provinces%20of%20South%20Africa.

7. Calvin, K., Dasgupta, D., Krinner, G., Mukherji, A., Thorne, P. W., Trisos, C., Romero, J., Aldunce, P., Barrett, K., Blanco, G., Cheung, W. W. L., Connors, S., Denton, F., Diongue-Niang, A., Dodman, D., Garschagen, M., Geden, O., Hayward, B., Jones, C., … Ha, M. (2023). IPCC, 2023: Climate Change 2023: Synthesis Report. Contribution of Working Groups I, II and III to the Sixth Assessment Report of the Intergovernmental Panel on Climate Change [Core Writing Team, H. Lee and J. Romero (eds.)]. *IPCC*, *Geneva*, *Switzerlan*(P*d*.. Arias, M. Bustamante, I. Elgizouli, G. Flato, M. Howden, C. Méndez-Vallejo, J. J. Pereira, R. Pichs-Madruga, S. K. Rose, Y. Saheb, R. Sánchez Rodríguez, D. Ürge-Vorsatz, C. Xiao, N. Yassaa, J. Romero, J. Kim, E. F. Haites, Y. Jung, R. Stavins, … C. Péan, Eds.). 10.59327/IPCC/AR6-9789291691647

8. Carlton, E. J., Woster, A. P., DeWitt, P., Goldstein, R. S., & Levy, K. (2016). A systematic review and meta-analysis of ambient temperature and diarrhoeal diseases. In International Journal of Epidemiology (Vol. 45, Issue 1, pp. 117–130). Oxford University Press. 10.1093/ije/dyv296

9. Centre for Research on the Epidemiology of Disasters (CRED). (2019). The human cost of disasters: An overview of the last 20 years *2000-*2.0 https://www.undrr.org/quick/50922

10. Chersich, M. F., Wright, C. Y., Venter, F., Rees, H., Scorgie, F., & Erasmus, B. (2018). Impacts of climate change on health and wellbeing in South Africa. In International Journal of Environmental Research and Public Health(Vol. 15, Issue 9). MDPI. 10.3390/ijerph15091884

11. Colston, J. M., Zaitchik, B. F., Badr, H. S., Burnett, E., Ali, S. A., Rayamajhi, A., Satter, S. M., Eibach, D., Krumkamp, R., May, J., Chilengi, R., Howard, L. M., Sow, S. O., Jahangir Hossain, M., Saha, D., Imran Nisar, M., Zaidi, A. K. M., Kanungo, S., Mandomando, I., … Kosek, M. N. (2022). Associations Between Eight Earth Observation-Derived Climate Variables and Enteropathogen Infection: An Independent Participant Data Meta-Analysis of Surveillance Studies With Broad Spectrum Nucleic Acid Diagnostics. GeoHealth, *6*(1). 10.1029/2021GH000452

12. Department of Environment Forestry and Fisheries Republic of South Africa. (2019). *National Climate Change Adaptation Strategy Republic of South Afri*.*ca* https://www.dffe.gov.za/sites/default/files/docs/nationalclimatechange_adaptationstrategy_ue10november2019.pdf

13. Department of Environmental Affairs and Tourism. (2004). *A National Climate Change Response Strategy for South Afric*.*a* https://unfccc.int/sites/default/files/sem_sup3_south_africa.pdf

14. Destoumieux-Garzón, D., Matthies-Wiesler, F., Bierne, N., Binot, A., Boissier, J., Devouge, A., Garric, J., Gruetzmacher, K., Grunau, C., Guégan, J. F., Hurtrez-Boussès, S., Huss, A., Morand, S., Palmer, C., Sarigiannis, D., Vermeulen, R., & Barouki, R. (2022). Getting out of crises: Environmental, social-ecological and evolutionary research is needed to avoid future risks of pandemics. In Environment International (Vol. 158). Elsevier Ltd. 10.1016/j.envint.2021.106915

15. Dickinson, N., Henriquez, F., Lynch, M., & Spencer, L. (2024). *Systematic review of the literature investigating emerging trends in Extreme Weather Events (EWEs) and infectious disease outbreaks in South Afric*.*a* https://www.crd.york.ac.uk/prospero/export_record_pdf.php

16. Dickinson, N., Spencer, L. H., Yang, S., Miller, C., Hursthouse, A., & Lynch, M. (2024). Extreme weather events in the UK and resulting public health outcomes. In MedRxiv . 10.1101/2024.11.25.24317884

17. Godsmark, C. N., Irlam, J., van der Merwe, F., New, M., & Rother, H. A. (2019). Priority focus areas for a sub-national response to climate change and health: A South African provincial case study. In Environment International (Vol. 122, pp. 31–51). Elsevier Ltd. 10.1016/j.envint.2018.11.035

18. Grobusch, L. C., & Grobusch, M. P. (2022). A hot topic at the environment–health nexus: investigating the impact of climate change on infectious diseases. In International Journal of Infectious Diseases (Vol. 116, pp. 7–9). Elsevier B.V. 10.1016/j.ijid.2021.12.350

19. Haines, A., Kovats, R. S., Campbell-Lendrum, D., & Corvalan, C. (2006). Climate change and human health: Impacts, vulnerability and public health. Public Health, 120(7), 585–596. 10.1016/j.puhe.2006.01.002

20. Hassell, J. M., Begon, M., Ward, M. J., & Fèvre, E. M. (2017). Urbanization and Disease Emergence: Dynamics at the Wildlife–Livestock–Human Interface. In Trends in Ecology and Evolutio(nVol. 32, Issue 1, pp. 55–67). Elsevier Ltd. 10.1016/j.tree.2016.09.012

21. Hunter, P. R. (2003). Climate change and waterborne and vector-borne disease. Journal of Applied Microbiology Symposium Supplement, 94(32). 10.1046/j.1365-2672.94.s1.5.x

22. Ikeda, T., Behera, S. K., Morioka, Y., Minakawa, N., Hashizume, M., Tsuzuki, A., Maharaj, R., & Kruger, P. (2017). Seasonally lagged effects of climatic factors on malaria incidence in South Africa. Scientific Reports, 7(1). 10.1038/s41598-017-02680-6

23. Ikeda, T., Kapwata, T., Behera, S. K., Minakawa, N., Hashizume, M., Sweijd, N., Mathee, A., & YaelWright, C. (2019). Climatic factors in relation to diarrhoea hospital admissions in rural Limpopo, South Africa. Atmosphere, 10(9). 10.3390/atmos10090522

24. Irlam, J., Reid, S., & Rother, H.-A. (2024). Education about planetary health and sustainable healthcare: A national Delphi panel assessment of its integration into health professions education in South Africa. *African Journal of Health Professions Educatio*, n16(1), e327. 10.7196/ajhpe.2024.v16i1.327

25. Kapuka, A., & Hlásny, T. (2021). Climate change impacts on ecosystems and adaptation options in nine countries in southern Africa: What do we know? Ecosphere, 12(12). 10.1002/ecs2.3860

26. Kapwata, T., Kunene, Z., Wernecke, B., Lange, S., Howard, G., Nijhawan, A., & Wright, C. Y. (2022). Applying a WASH Risk Assessment Tool in a Rural South African Setting to Identify Risks and Opportunities for Climate Resilient Communities. *International Journal of Environmental Research and Public Healt*,h19(5). 10.3390/ijerph19052664

27. Kapwata, T., Wright, C. Y., du Preez, D. J., Kunene, Z., Mathee, A., Ikeda, T., Landman, W., Maharaj, R., Sweijd, N., Minakawa, N., & Blesic, S. (2021). Exploring rural hospital admissions for diarrhoeal disease, malaria, pneumonia, and asthma in relation to temperature, rainfall and air pollution using wavelet transform analysis. *Science of the Total Environmen*, t791. 10.1016/j.scitotenv.2021.148307

28. Khine, M. M., & Langkulsen, U. (2023). The Implications of Climate Change on Health among Vulnerable Populations in South Africa: A Systematic Review. In International Journal of Environmental Research and Public Health(Vol. 20, Issue 4). MDPI. 10.3390/ijerph20043425

29. Kim, H., Chae, Y., & Kim, S. (2021). Climate change, extreme weather events, and human infectious diseases: A systematic review. . International Journal of Envrionmental Research and Public Health, 18(9), 4488.

30. Kitole, F. A., Mbukwa, J. N., Tibamanya, F. Y., & Sesabo, J. K. (2024). Climate change, food security, and diarrhoea prevalence nexus in Tanzania. Humanities and Social Sciences Communication,s 11(1). 10.1057/s41599-024-02875-z

31. Kovats, S., & Akhtar, R. (2008). Climate, climate change and human health in Asian cities. Environment and Urbanization, 20(1), 165–175. 10.1177/0956247808089154

32. Lee, T. T., Dalvie, M. A., Röösli, M., Merten, S., Kwiatkowski, M., Mahomed, H., Sweijd, N., & Cissé, G. (2023). Understanding diarrhoeal diseases in response to climate variability and drought in Cape Town, South Africa: a mixed methods approach. Infectious Diseases of Povert,y*12*(1). 10.1186/s40249-023-01127-7

33. Lequechane, J. D., Mahumane, A., Chale, F., Nhabomba, C., Salomão, C., Lameira, C., Chicumbe, S., & Semá Baltazar, C. (2020). Mozambique’s response to cyclone Idai: How collaboration and surveillance with water, sanitation and hygiene (WASH) interventions were used to control a cholera epidemic. *Infectious Diseases of Povert*,y9(1). 10.1186/s40249-020-00692-5

34. Liao, H., Lyon, C. J., Ying, B., & Hu, T. (2024). Climate change, its impact on emerging infectious diseases and new technologies to combat the challenge. In Emerging Microbes and Infections (Vol. 13, Issue 1). Taylor and Francis Ltd. 10.1080/22221751.2024.2356143

35. Luyt, C. D., Tandlich, R., Muller, W. J., & Wilhelmi, B. S. (2012). Microbial Monitoring of Surface Water in South Africa: An Overview. In International Journal of Environmental Research and Public Health (Vol. 9, Issue 8, pp. 2669–2693). MDPI. 10.3390/ijerph9082669

36. Lynch, M., Harris, F., Ierna, M., Mahomed, O., Henriquez-Mui, F., Gebreslasie, M., Ndzi, D., Viriri, S., Shakir, M. Z., Dickinson, N., Miller, C., Nadesan-Reddy, N., Nkwanyana, F., Spencer, L. H., & Naidoo, S. (2025). Warning system for Extreme weather events, Awareness Technology for Healthcare, Equitable delivery, and Resilience (WEATHER) Project: A mixed methods research study protocol. 10.1101/2025.01.14.25320537

37. MacDonald, A. J., & Mordecai, E. A. (2019). Amazon deforestation drives malaria transmission, and malaria burden reduces forest clearing. *Proceedings of the National Academy of Sciences of the United States of Americ*,a116(44), 22212–22218. 10.1073/pnas.1905315116

38. Martineau, P., Behera, S., Nonaka, M., Jayanthi, R., Ikeda, T., Minakawa, N., Kruger, P., & Mabunda, Q. (2022). Predicting malaria outbreaks from sea surface temperature variability up to 9 months ahead in Limpopo, South Africa, using machine learning. Frontiers in Public Healt.h 10.3389/fpubh.2022.962377

39. McMichael, A. J. (2015). Extreme weather events and infectious disease outbreaks. Virulence, 6(6), 543–547. 10.4161/21505594.2014.975022

40. Meurens, F., Dunoyer, C., Fourichon, C., Gerdts, V., Haddad, N., Kortekaas, J., Lewandowska, M., Monchatre-Leroy, E., Summerfield, A., Wichgers Schreur, P. J., van der Poel, W. H. M., & Zhu, J. (2021). Animal board invited review: Risks of zoonotic disease emergence at the interface of wildlife and livestock systems. In Animal (Vol. 15, Issue 6). Elsevier B.V. 10.1016/j.animal.2021.100241

41. Meyer, C. A. (2024). The Extent of the Inclusion and Consideration of Extreme Climate Events and Health in South African Policies; The Case of eThekw. Ini https://wiredspace.wits.ac.za/items/aafb0e68-afa4-4817-8a9d-1476abacf68b

42. Moher, D., Liberati, A., Tetzlaff, J., Altman, D. G., Altman, D., Antes, G., Atkins, D., Barbour, V., Barrowman, N., Berlin, J. A., Clark, J., Clarke, M., Cook, D., D’Amico, R., Deeks, J. J., Devereaux, P. J., Dickersin, K., Egger, M., Ernst, E., … Tugwell, P. (2009). Preferred reporting items for systematic reviews and meta-analyses: The PRISMA statement. PLoS Medicine, 7(9), 889–896. 10.3736/jcim20090918

43. Moola, S., Munn, Z., Tufanaru, C., Aromataris, E., Sears, K., Sfetcu, R., Currie, M., Qureshi, R., Mattis, P., Lisy, K., & Mu, P.-F. (2017). Checklist for Cohort Studies. Joanna Briggs Institute Reviewer’s Manual, 1–7. https://joannabriggs.org/ebp/critical_appraisal_tools

44. Moyo, E., Mhango, M., Moyo, P., Dzinamarira, T., Chitungo, I., & Murewanhema, G. (2023). Emerging infectious disease outbreaks in Sub-Saharan Africa: Learning from the past and present to be better prepared for future outbreaks. In Frontiers in Public Health(Vol. 11). Frontiers Media S.A. 10.3389/fpubh.2023.1049986

45. Munn, Z., Barker, T., Moola, S., Tufanaru, C., Stern, C., McArthur, A., Stephenson, M., & Aromataris, E. (2021). Methodological quality of case series studi.eJsBI Evidence Synthesis. doi: 10.11124/JBISRIR-D-19-00099

46. Ngcobo, N., Mzinyane, B., & Zibane, S. (2023). Responding to Concurrent Disasters: Lessons Learnt by Social Work Academics Engaging with Flood Survivors during a COVID-19 Pandemic, in South African Townships. https://www.mdpi.com/books/edition/7335/article/8119-responding-to-concurrent-disasters-lessons-learnt-by-social-work-academics-engaging-with-flood

47. Nyam, Y. S., Modiba, N. T. S., Ojo, T. O., Ogundeji, A. A., Okolie, C. C., & Selelo, O. T. (2024). Analysis of the perceptions of flood and effect of adoption of adaptation strategies on income of informal settlements of Mamelodi in South Africa. Climate Services, 34. 10.1016/j.cliser.2024.100468

48. Olanrewaju, C., & Reddy, M. (2023). Vulnerability of Informal Settlements to Flood Disasters: A Review of Public Health Implication

49. Orievulu, K., Ayeb-Karlsson, S., Ngwenya, N., Ngema, S., McGregor, H., Adeagbo, O., Siedner, M. J., Hanekom, W., Kniveton, D., Seeley, J., & Iwuji, C. (2022). Economic, social and demographic impacts of drought on treatment adherence among people living with HIV in rural South Africa: A qualitative analysis. Climate Risk Management, 36. 10.1016/j.crm.2022.100423

50. Page, M., McKenzie, J., Bossuyt, P., Boutron, I., Hoffmann, T., & Mulrow, C. (2021). The PRISMA 2020 statement: an updated guideline for reporting systematic reviews. BMJ, 10(89). doi10.1186/s13643-021-01626-4

51. Potgieter, N., De Beer, M. C., Taylor, M. B., & Steele, A. D. (2010). Prevalence and diversity of rotavirus strains in children with acute diarrhea from rural communities in the Limpopo Province, South Africa, from 1998 to 2000. Journal of Infectious Disease,s *202*(SUPPL. 1). 10.1086/653561

52. Raza, M., Fatima, A., Habiba, U., & Shah, H. H. (2023). Public health implications of severe floods in Pakistan: assessing the devastating impact on health and the economy. Frontiers in Environmental Science, 11. 10.3389/fenvs.2023.1091998

53. Saatchi, M., Khankeh, H. R., Shojafard, J., Barzanji, A., Ranjbar, M., Nazari, N., Mahmodi, M. A., Ahmadi, S., & Farrokhi, M. (2024). Communicable diseases outbreaks after natural disasters: A systematic scoping review for incidence, risk factors and recommendations. In Progress in Disaster Science (Vol. 23). Elsevier Ltd. 10.1016/j.pdisas.2024.100334

54. Semenza, J., & Paz, S. (2021). Climate change and infectious disease in Europe: Impact, projections and adaptation. The Lancet Regional Health - Europe. 10.1016/j.lanepe.2021.100203

55. Shackleton, D., Memon, F. A., Nichols, G., Phalkey, R., & Chen, A. S. (2024). Mechanisms of cholera transmission via environment in India and Bangladesh: State of the science review. In Reviews on Environmental Health(Vol. 39, Issue 2, pp. 313–329). Walter de Gruyter GmbH. 10.1515/reveh-2022-0201

56. World Organisation for Animal Health. (2015). 21st Conference of the OIE Regional Commission for Africa. https://rr-africa.woah.org/en/news/21st-conference-of-the-reg-commission-rabat/

57. Wright, C. Y., Kapwata, T., du Preez, D. J., Wernecke, B., Garland, R. M., Nkosi, V., Landman, W. A., Dyson, L., & Norval, M. (2021). Major climate change-induced risks to human health in South Africa. Environmental Research, 196. 10.1016/j.envres.2021.110973

58. Yuan, J., Cao, Z., Ma, J., Li, Y., Qiu, Y., & Duan, H. (2024). Influence of climate extremes on long-term changes in cyanobacterial blooms in a eutrophic and shallow lake. Science of the Total Environment, 939. 10.1016/j.scitotenv.2024.173601

