## Supplementary File 1 for "A systematic review investigating emerging trends between Extreme Weather Events (EWEs) and infectious disease outbreaks in South Africa"

**NIHR WEATHER SR: Search Strategy**

**Search Date:** 11/06/2024

**Dates searched:** Jan 2014 to June 2024

**EBSCO Platform (12 databases) -** **208** (automatic deduplication - 162 into Endnote)

(All databases selected minus eBook collection, PsychBooks, Regional Business News, eBook Open Access Collection and Sage - Transforming Nursing Practice (eBook Sub).)

Abstract search selected.

AB "South Africa" AND ("Extreme Weather Events" OR "Meteorological conditions" OR "Climate Change" OR "Global Warming" OR "Natural Disasters" OR Flood* OR Storms OR "Adverse weather" OR "Climate events" ) AND ( “New conditions” OR “emerging conditions” OR “Emerging trends” OR disease OR Illness OR “disease risk” OR health OR “infectious disease” OR outbreak OR gastrointestinal OR water-borne OR waterborne OR pathogen* OR bacteria OR virus OR amoeb* OR parasit* OR vector OR “E. coli” OR Pseudomonas OR “Vibrio cholerae” OR cholera OR Acanthamoeba OR Giardia OR Schistosomiasis OR cryptosporidium OR Contamination OR "Environmental exposure" OR Malaria)

**Web of Science - 474**

**Topic search selected.**

**((TS=(“South Africa”)) AND TS=(“New conditions” OR “emerging conditions” OR “Emerging trends” OR “disease” OR “Illness” OR “disease risk” OR "health" OR “infectious disease” OR “outbreak” OR “gastrointestinal” OR “water-borne” OR "waterborne" OR “pathogen*” OR “bacteria” OR “virus” OR “amoeb*” OR “parasit*” OR “vector” OR “E. coli” OR “Pseudomonas” OR “Vibrio cholerae” OR “cholera” OR “Acanthamoeba” OR “Giardia” OR “Schistosomiasis” OR “cryptosporidium” OR "Contamination" OR "Environmental exposure" OR “Malaria” )) AND TS=("Extreme Weather Events" OR "Meteorological conditions" OR "Climate Change" OR "Global Warming" OR "Natural Disasters" OR "Flood*" OR "Storms" OR "Adverse weather" OR "Climate events" )**

**Science direct - 366**

‘Title, abstract or author-specified keywords’ selected.

Filtered to research papers and review papers.

Search terms adjusted as only 8 Boolean operators allowed:

**“South Africa”** in ‘find articles with these terms’ **and (Disease OR “human health” OR “infectious disease” OR outbreak OR pathogen) AND ("Extreme Weather Events" OR "Climate Change")**

**Cochrane – 8 (trials)**

Reviews and trials selected.

**"**South Africa" in Title Abstract Keyword AND "Extreme Weather Events" OR "Meteorological conditions" OR "Climate Change" OR "Global Warming" OR "Natural Disasters" OR Flood* OR Storms OR "Adverse weather" OR "Climate events" in Title Abstract Keyword AND “New conditions” OR “emerging conditions” OR “Emerging trends” OR disease OR Illness OR “disease risk” OR health OR “infectious disease” OR outbreak OR gastrointestinal OR “water-borne” OR waterborne OR pathogen* OR bacteria OR virus OR amoeb* OR parasit* OR vector OR “E. coli” OR Pseudomonas OR “Vibrio cholerae” OR cholera OR Acanthamoeba OR Giardia OR Schistosomiasis OR cryptosporidium OR Contamination OR "Environmental exposure" OR Malaria in Title Abstract Keyword - with Cochrane Library publication date Between Jan 2014 and Jun 11th 2024, in Cochrane Reviews (Word variations have been searched)

**Total results:**

8+208+366+474 = 1056

EBSCO removed 46 duplicates

Endnote removed 75 duplicates

Rayyan removed a further 20 duplicates

Total duplicates = 141

1056 – 141 = **915**

**Filtering:**

**Inclusion criteria:** All papers relating to extreme weather events impact on infectious disease in South Africa, written in the English language from 2014 to the present 2024.

**Exclusion criteria:** papers not related to extreme weather events influence on infectious disease outbreaks in SA.

**Stage 1 Filtering:**

915 papers filtered by title and abstract by two authors - 46 papers remaining.

**Stage 2 Filtering:**

Full text screening by two reviewers, followed by further discussion between four of the authors - 12 papers included for data extraction.

**Hand Searching:**

2 additional papers identified through reference list screening: Ikeda et al. (2019) and Ikeda et al. (2017).

**Final Number of Papers: 14**
