## Supplementary File 2 for "A systematic review investigating emerging trends between Extreme Weather Events (EWEs) and infectious disease outbreaks in South Africa"

### Data extraction form for WEATHER Systematic Review

| **Study Details** | **Intervention** | **Study type** | **Outcomes** | **Main findings** |
| --- | --- | --- | --- | --- |
| **Author:** Godsmark *et al.*  **Year:** 2019  **DOI:**  **Title:**  **Country:** South Africa  **Aim:** To highlight priority focus areas for a sub-national government response to health and climate change, using the Western Cape (WC) province of South Africa as a case study. | **Intervention:** No intervention  **Dates of data collection:** The main review documents were published between 2011 and 2015.  **Population and sample size:** N/A  **Setting:** Western Cape (WC), South Africa.  **Delivery mode (e.g., remotely online, in person):** - N/A  **Intervention deliverers:** No intervention  **Timing and duration:** No intervention  **Intervention description:** No intervention | **Study type:** Review of priority areas.  **Review papers:** Six main review documents were chosen and other literature added to answer questions about the priority areas.  **Length of follow-up:** N/A | **Outcome/s of interest:**  Infectious diseases including:  Ebola  Avian Influenza  TB  Hepatitis E  Hepatitis General  Meningococcal meningitis  Colds and flu  Measles  Pneumonia  Hand, foot and mouth disease (PSG)  Cholera  Typhoid  Diarrhoeal diseases  Legionnaires disease (PSG)  Hepatitis A. | **Main finding:** Western Cape government priority focus areas requiring further research on health risk factors include: population, migration and environmental refugees, land use change, violence and human conflict and vulnerable groups. WC government priority focus areas for further research on health impacts include: mental ill-health, non-communicable diseases, injuries, poisonings (e.g. pesticides), food and nutrition insecurity-related diseases, water and food-borne diseases and reproductive health. These areas are currently under addressed, or not addressed at all, in the current provincial climate change strategy.  **Additional finding:**  Water-borne diseases are highly susceptible to increases in temperature and also surge in times of drought due to poor sanitation and hygiene practices, as well as reduced water quality). Relationships have been established between increased temperature and/or extreme weather events such as flooding, heavy rainfall, drought, El Niño Southern Oscillation and hurricanes; and hepatitis, rotavirus, norovirus, enterovirus, cholera, giardia, typhoid and legionnaires disease. For coastal regions, an increase in harmful algal blooms and increased ocean temperatures have been correlated with outbreaks of cholera as the bacteria is transmitted to humans through the consumption of raw shellfish). Water-borne diseases and diarrhoea often result in severe dehydration, which can be fatal. |
| **Author:** Kapwata *et al.*  **Year:** 2022  **DOI:**  **Title:**  **Country:** South Africa  **Aim:** To conduct a risk assessment of two climate-related variables (i.e., temperature and rainfall) and associated water, sanitation and hygiene (WASH)-related exposures and vulnerabilities for people living in Mopani District, Limpopo province, South Africa. | **Intervention:** No intervention  **Dates of data collection:** Between 1958 and 2017.  **Population and sample size:** People living in Mopani District, Limpopo province, South Africa.  **Setting:** South Africa  **Delivery mode (e.g., remotely online, in person):** N/A  **Intervention deliverers:** N/A  **Timing and duration:** N/A  **Intervention description:** N/A | **Study type:** Review of climate change policies and adaptation plans for the water supply and sanitation sectors of Mopani District.  **Length of follow-up:** N/A | **Outcome/s of interest:**  Water quality. | **Main finding:**  The Mopani District is well known for experiencing heavy rain and flooding and adequate data exist to support this classification.  In the Mopani District, 13% of More than a third of households were exposed to microorganisms that can cause diseases and that may come via piped water provided by the municipality.  **Additional finding:** Data from the South African 2015/2016 District Health Barometer showed that the burden of diarrhoeal diseases is high in Mopani. The District reported a case fatality rate due to diarrhoea of 4%, which is above the national target of 3%. This was classified as high exposure. |
| **Author:** Khine and Langkulsen  **Year:** 2023  **DOI:**  **Title:**  **Country:** South Africa  **Aim:** To identify the role of climate change in increasing multidimensional inequalities among vulnerable populations and analyse the strengths and limitations of South Africa’s National Climate Change Adaptation Strategy. | **Intervention:** No intervention  **Dates of data collection:** Literature from 2014-2022  **Population and sample size:** 854 identified sources, and 24 included in the review.  **Setting:** South Africa  **Delivery mode (e.g., remotely online, in person):** N/A  **Intervention deliverers:** N/A  **Timing and duration:** N/A  **Intervention description:** N/A | **Study type:** Systematic review  **Length of follow-up:** N/A | **Outcome/s of interest:**  Flooding and the consequent health risks.  Water availability  Sanitation | **Main finding:** In South Africa, a lack of a robust database system generating climate information and assessing the impacts hinders effective action. This makes it difficult for decision makers to identify sufficient resources for climate change adaption.  For an inclusive and sustainable reduction in inequalities and vulnerabilities to the impact of climate change, community-based health and social services should be enhanced among vulnerable populations.  **Additional finding:** Floods, storms and droughts are likely to worsen in South Africa.  Climate change has a significant impact on water resources. A lack of access to water for daily use such as cooking and drinking, poses a serious threat of epidemics and other waterborne diseases. |
| **Author:** Abrams *et al*.  **Year:** 2021  **DOI:** <https://doi.org/10.3390/w13202810>  **Title:** Water, Sanitation, and Hygiene Vulnerability among Rural Areas and Small Towns in South Africa: Exploring the Role of Climate Change, Marginalization, and Inequality.  **Country:** South Africa  **Aim:** To explore the needs, barriers, and vulnerabilities with respect access to water, sanitation, and hygiene (WASH) in rural areas and small towns in South Africa—using two case studies to explore climate risk and vulnerability assessment (CRVA) in a rural village and urban municipality | **Intervention:**  No intervention- WASH-focused case studies  **Dates of data collection:**  17 and 18 May 2018  **Population and sample size:** 80 people interviewed 5 times  **Setting:** South Africa  **Delivery mode (e.g., remotely online, in person):**  observations, in-depth interviews, surveys, focus groups, desk-based research, and a wide range of participatory exercises and activities  **Intervention deliverers:**  **Timing and duration:**  study was conducted from 2013 through 2016  **Intervention description:** | **Study type:**  Observation study plus qualitative interviews with key stakeholders  **Length of follow-up:**  4 years of project | **Outcome/s of interest:**  CVRA WASH case studies  background picture for the cases  1. Historic context—including land ownership/dispossession and resource access;  2.Climate, extreme weather events, natural resources context;  3. Socio-economic profile.  The analysis included the following:  1. WASH specific analysis;  2.Catchment vulnerability assessment;  3. Source vulnerability assessment;  4.WASH management and community engagement capacity/vulnerability assessment;  5.WASH supply chain and supporting infrastructure vulnerabilities and capacity assessment. | **Main finding:**  WASH services in rural and small towns in South Africa are influenced by several factors—history, natural elements, and socio-economic aspects are intertwined and influence WASH vulnerability.  Findings indicate the importance of fieldwork and engaging outside the academic sphere.  Quick assessments can provide some information, but often miss out on the finer details.  During flooding events, water accessibility is challenged by issues of access to spare parts; therefore to ensure resilience in a flooding event is to retain ample stocks of spare parts for all essential WASHD infrastructure.  Trust is established between management and local communities  **Additional finding:**  local organizations take over water delivery, there should be mechanisms in place (via policy and funding) to support such efforts. |
| **Author:** Orievulu *et al.*  **Year:** 2021  **DOI:** <https://doi.org/10.1016/j.crm.2022.100423>  **Title:**  **Country:** South Africa  **Aim:**  To examine how drought’s multi-dimensional effects on interlinked socioeconomic and demographic factors potentially heightened challenges to optimal HIV care among PLHIV. The project aims to demonstrate the multiple pathways by which the 2015 drought intensified existing challenges associated with HIV treatment adherence. | **Intervention:**  **Dates of data collection:**  August 2019 and June 2020  **Population and sample size:** 27 individuals interviewed  **Setting:** KZN South Africa  **Delivery mode (e.g., remotely online, in person):**  systems approach, embedded in the qualitative methodology, to explore, and possibly explain, pathways by which drought contributes to failing HIV care among PLHIV.  **Intervention deliverers:**  **Timing and duration:**  N/A  **Intervention description:** N/A | **Study type:**  Face-to-face in-depth Interviews (IDI) and telephonic in-depth interviews (TIDI)  **Length of follow-up:**  1 year | **Outcome/s of interest:**  The goal is to show the complexities underlying how the experience of the 2015 drought potentially contributed to an exacerbation of pre-existing contextual vulnerabilities and precarious livelihoods with potentially challenging effects on HIV care.  To understand that taking a systems approach would allow exploration of possible interconnections by recognising and identifying the intermediate factors triggered or exacerbated by drought | **Main finding:**  Drought- enforced soil water depletion, dried-up rivers, and dams terminated in a continuum of events such as loss of livestock, reduced agricultural production, and insufficient access to water and food which was understood to indirectly have a negative impact on HIV treatment adherence.  This led to disruptions in incomes, livelihoods and food systems, increased risk to general health, forced mobility and exacerbation of contextual vulnerabilities linked to poverty and unemployment.  **Additional finding:**  Taking a systems approach highlighted the complex pathways of plausible networks of impacts from drought through varying socioeconomic factors, exacerbating longstanding contextual precarity, and ultimately challenging HIV care utilisation. This facilitated depiction of the multidimensional relationships between climate change, especially drought, and poor HIV care outcomes through the prism of contextual vulnerabilities is vital for shaping policy interventions. |
| **Author:** Amegah *et al.*  **Year:** 2016  **DOI:**  **Title:**  **Country:** Sub-Saharan Africa, including SA  **Aim:** To systematically review all studies investigating temperature variability and non-vector borne morbidity and mortality in SSA to establish the state and quality of available evidence, identify  gaps in knowledge, and propose future research priorities | **Intervention:** No intervention  **Dates of data collection:** The main review documents were published between 2004 and 2014.  **Population and sample size:** N/A  **Setting:** SS Africa  **Delivery mode (e.g., remotely online, in person):** -  **Intervention deliverers:** No intervention  **Timing and duration:** No intervention  **Intervention description:** No intervention | **Study type:** systematic review  **Review papers:** 23 in the inclusion criteria of climate OR climatic OR weather OR temperature} AND {mortality OR death* OR morbidity OR illness* OR disease* OR sickness* OR  infection* OR malnutrition OR undernutrition OR diarrhoea OR cholera}  AND africa.  **Length of follow-up:** N/A | **Outcome/s of interest:**  Temperature associated Infectious diseases including:  Ebola  Cholera  Diarrhoeal diseases  Respiratory disease  Non-communicable challenges:  Malnutrition  Cardiovascular disease  Skin disease | **Main finding:** temperature exposure may contribute to disease and mortality burden in SSA with an increase in Cholera, CVD, and diarrhoeal disease in higher temperatures. Ebola and respiratory diseases in lower temperatures. Undernutrition in children is higher in elevated temperatures. However, it must be acknowledged that the evidence-base is weakened by the limited number of studies and methodological limitations of studies.  **Additional finding:** There are limited meteorological stations, poor health surveillance and record keeping that contribute to the challenges of climate-health related research. Investment is required. Profiling socio-economic differentials should also be considered to highlight any exacerbation of temperature-health relationships. |
| **Author:** Chersich *et al.*  **Year:**  **DOI:**  **Title:**  **Country:**  **Aim:** | **Intervention:** No intervention  **Dates of data collection:** The main review documents were published between 2010 and 2017.  **Population and sample size:** N/A  **Setting:** South Africa  **Delivery mode (e.g., remotely online, in person):** -  **Intervention deliverers:** No intervention  **Timing and duration:** No intervention  **Intervention description:** No intervention | **Study type:** systematic review  **Review papers:**  **34 papers identified from** ((“South Africa”[MeSH]) OR (“South Africa”[Title/Abstract]) OR (“Southern Africa*“[Title/Abstract])))  AND “last 10 years”[PDat])) AND (((“global warming”[Title/Abstract] OR “global warming”[MeSH]  OR climatic*[Title/Abstract] OR “climate change”[Title/Abstract] OR “climate change”[MeSH] OR  “Desert Climate”[MeSH] OR “El Nino-Southern Oscillation”[MeSH] OR Microclimate[MeSH] OR  “Tropical Climate”[MeSH]))  **Length of follow up:** NA | **Outcome/s of interest:**  Infectious disease  Malaria  Dengue  Zika  Avian Influenza  Cholera  Schistosomiasis  Mental Health  Gender-based violence  Suicide  Food insecurity | **Main finding:**  very few studies reviewed used empirical data from health  services to analyse the impact of climate change. More effective use of surveillance and research data are required to monitor climate change impacts on human health in South Africa. Relevant studies focused on infectious disease, but mainly malaria.  **Additional finding:**  Consequences are direct and indirect  Direct mainly on infectious diseases as climate change has direct effect on pathogen numbers and ability to colonise food and water. Indirect effects on human health mainly on mental health and stress. |
| **Author:** Colston *et al.*  **Year:** 2022  **DOI:**  **Title:** Associations Between Eight Earth Observation-Derived Climate Variables and Enteropathogen Infection: An Independent Participant Data Meta-Analysis of Surveillance Studies With Broad Spectrum Nucleic Acid Diagnostics.  **Country:** South Africa  **Aim:** to pool data from comparable studies in multiple locations across diverse geographical areas and climate zones and match them with coincident Earth observation-derived data to model the associations between eight hydrometeorological exposures (precipitation, runoff volume, humidity, soil moisture, solar radiation, air pressure, temperature, and wind speed) and infection status for 10 common, high-burden enteric pathogens ascertained in young children (adenovirus, astrovirus, norovirus, rotavirus, sapovirus, Campylobacter, ETEC, Shigella, Cryptosporidium, and Giardia). The research question to be assessed was whether different combinations of rainfall, ambient temperature, atmospheric humidity and pressure and other parameters impact the risk of enteric infections independently of each other and of seasonality, and at magnitudes and in directions that differ by pathogen species, which are differentially resistant to environmental conditions such as dryness, ultraviolet light, and in their probability of transmission via aerosol | **Intervention:** No intervention  **Dates of data collection:** The main review documents were published between 2012 and 2022  **Population and sample size:** Studies discussed were at global level and included cases of diarrhoea from different types of populations, mainly children, women in pregnancy  **Setting:** global, many countries including South Africa  2 studies included SA, while a further 6 included sites in SSA.  **Delivery mode (e.g., remotely online, in person):** -  **Intervention deliverers:** No intervention  **Timing and duration:** No intervention  **Intervention description:** No intervention | **Study type:** meta-analysis with new data generation  **Review papers:** 15 publications/studies that adhere to inclusion criteria  Some sites, including South Africa did not test for all pathogens  **Length of follow up:** N/A | **Outcome/s of interest:**  Diarrhoeal disease caused by different viruses, bacteria and protozoa  The enteropathogens include:  Viruses:  Adenovirures, rotaviruses, astroviruses, sapoviruses  Bacteria:  Campylobacter, ETEC, Shigella/EIEC  Protozoa:  Cryptosporidium, Giardia | **Main finding:**  Four main hypothesized mechanisms by which hydrometeorological conditions impact short-term risk of enteropathogen transmission include 1. waterborne dispersal 2. airborne dispersal  3. survival on surfaces and 4. fomites.  Enteric viruses during periods when precipitation fell below the local average, with the exception of rotavirus which was, conversely, the only virus for which risk increased with heavier surface runoff. The only single pathogen that appeared to increase in prevalence with both low rainfall and heavy runoff was ETEC.  Protozoal infections correlate with increased rainfall  **Additional finding:**  Lag in infection post rainfall. There are strong associations of soil moisture, humidity and temperature with almost all pathogen taxa.  ‘ Rising global temperatures may lead to decreases in adenovirus and rotavirus burden, and increases in the prevalence of enteric protozoa and bacteria, most notably ETEC, while the **impacts of greater precipitation variability due to climate change on diarrhea-causing pathogens are less certain** and likely to be highly species- and location-specific.’ |
| **Author:** Wright *et al.*  **Year:** 2021  **DOI:**  **Title:**  **Country:** South Africa  **Aim:** To describe the major climatic changes facing South Africa and how they can impact on human health | **Intervention:** No intervention  **Dates of data collection: ?**  **Population and sample size:** N/A  **Setting:** South Africa.  **Delivery mode (e.g., remotely online, in person):** -  **Intervention deliverers:** No intervention  **Timing and duration:** No intervention  **Intervention description:** No intervention | **Study type:** Narrative review of priority areas.  **Review papers:**  Does not tell us anything about the papers included in the review.  **Length of follow-up:** N/A  **Limitations:**  Narrative review only | **Outcome/s of interest:**  Human health risks due to climate change and EWEs. | **Main finding:**  HIV – increases associated with vulnerability – access to medical care, drug supplies, non-adherence with treatment regimes. Increased risk of mother-child transmission in hot weather.  Malaria and other mosquito or tick borne infections– highest in wet, summer months.  Schistosomiasis most problematic in hot, wet seasons.  Extreme rainfall events – flooding events associated with cholera outbreak in Johannesburg.  Extreme rain increases prevalence of vector borne diseases  Higher rainfall led to higher diarrhoeal disease in under 5s in Limpopo province.  Surge in mosquito numbers after droughts, leading to malaria outbreaks when the first rains fall.  **Additional finding:**  Heat related effects are linked with future sustained increased temperatures, rather than EWEs. Mosquito borne, tick borne and schistosomiasis all predicted to spread to new geographically diverse areas due to increasingly favourable conditions (hotter and wetter) for the infectious agents. |
| **Author:** Abdullahi, Nitschke and Sweijd.  **Year:** 2022  **DOI:** https://doi. Org/10.1371/journal.pone.0262008  **Title:** Predicting diarrhoea outbreaks with climate change.  **Country:** South Africa  **Aim:**  To ascertain the suitability of various ML methods given various climate factors and synthetic (generative) training data for accurately predicting diarrhoea outbreaks. Specifically, the study aims to elucidate what type of ML method is most appropriate when coupled with specific training and test data-sets (that is, specific climate variables, data-sparseness, data-noise and synthetic data compliment), in order to optimise prediction efficacy. | **Intervention:** Compared **t**ask performance of 3 Machine Learning methods (CNNs, LSTMs and SVMs)  **Dates of data collection:** Used 10 years of records. 2008 - 2018  **Population and sample size:** 10 years of sales of loperamide as a proxy for diarrhoea cases.  Max and min temp, air temp, specific humidity, potential evaporation rate, precipitation rate, surface pressure and wind velocity climate factors obtained from the National centres for Atmoshpheric Research and Atmospheric Prediction  **Setting:** This study focused on the nine South African Provinces which are: Western Cape, Eastern Cape, Northern Cape, North West, Free State, Limpopo, KwaZulu Natal, Gauteng, and Mpumalanga.  **Delivery mode (e.g., remotely online, in person):** -  **Intervention deliverers:** No intervention  **Timing and duration:** No intervention  **Intervention description:** No intervention | **Study type:** Modelling study – comparing efficacy of 3 ML methods to predict daily diarrhoea cases in SA by incorporating climate information.  **Review papers: NA**  **Length of follow-up:** N/A | **Outcome/s of interest:**  Accuracy of ML methods to predict diarrhoea in various climate scenarios. | **Main finding:**  All three ML models were appropriate for predicting daily diarrhoea cases with selected climate variables. Data augmentation (AI) improved accuracy by 30%.  The most influential climate variables for predicting diarrhoea outbreak in SA were precipitation, humidity, evaporation and temperature.  **Additional finding:**  Suggest that taking other human and environmental factors that contribute to spread of infections disease into account may improve accuracy of future prediction models. |
| **Author:** Lee *et al.*  **Year:** 2023  **DOI:** https://doi.org/10.1186/s40249-023-01127-7  **Title:** Understanding diarrhoeal diseases in response to climate variability and drought in Cape Town, South Africa: a mixed methods approach.  **Country:** South Africa  **Aim:** to gain a more holistic understanding of the relationship between diarrhoea in young children and climate variability in a system stressed by water scarcity  Focus on diarrhoeal disease during drought rather than flooding. | **Intervention:** No intervention  **Dates of data collection:** 2012 - 2019  **Population and sample size:** diarrhoeal disease with dehydration in children <5, during the hot dry season (Nov – May)  monthly case count of diarrhoea with dehydration in under 5s presenting at primary healthcare facilities in Western Cape from 2010 to 2019.  Weather data on max and min temperature, precipitation and relative humidity was collected from SA Weather Service.  Key stakeholder interviews conducted with health/environment personnel. Purposive and snowball sampling.  **Setting:** Western Cape (WC), South Africa.  **Delivery mode (e.g., remotely online, in person):** -  **Intervention deliverers:** No intervention  **Timing and duration:**  **Intervention description:** No intervention | **Study type:** Mixed methods.  Negative binomial regression model developed to understand relationships between diarrhoeal incidence in the dry season, and climate factors.  14 semi-structured interviews with health and environmental workers to understand the reasons for observed trends. Analysed using the Framework method.  **Review papers:**  **Length of follow-up:** N/A | **Outcome/s of interest:**  Diarrhoeal disease with dehydration incidence, and its relationship with temperature, precipitation and relative humidity. | **Main finding:**  Incidence of diarrhoea with dehydration cases has fallen significantly in the period 2010-2019. A 64.7% reduction (95% CI: 5.5-7.2%, with no significant increase over the period of severe drought.  Interviews supported the quantitative data, with stakeholders reporting an obvious drop in diarrhoea, and attributing this to consistent public health interventions and messaging.    Max temp and rel. humidity are significantly positively associated with diarrhoea with dehydration, IRRs 1.074 (95% CI: 1.045 – 1.103) and 1.026 (95% CI:1.017 – 1.035) respectively.  This signifies an increase in diarrhoea with dehydration of 7% for each 1 degree increase in temp, and 3% for each 1% increase in rel. humidity during the diarrhoeal surge season    **Additional finding:**  Decrease in mortality and morbidity attributed to a national immunization programme for rotavirus, greater awareness of WaSH practices, improved standards of living, and wider delivery of health services.  Drought is a concern due to having water for keeping clean and safe drinking, not only at home, but in the hospital and clinic setting.  The study did not find an association between rainfall and diarrhoea, though they investigated climate effects at an aggregated level. It is likely that on a daily scale, the effect may be more noticeable. |
| **Author:** Kapwata *et al.*  **Year:** 2021  **DOI:** https://doi.org/10.1016/j.scitotenv.2021.148307.  **Title:** Exploring rural hospital admissions for diarrhoeal disease, malaria, pneumonia, and asthma in relation to temperature, rainfall and air pollution using wavelet transform analysis.  **Country:** South Africa  **Aim:** to investigate time series of daily admissions from two public hospitals in Limpopo province in South Africa with climate variability and air quality. | **Intervention:** No intervention  **Dates of data collection:** 2011 - 2017  **Population and sample size:**  2557 data points  **Setting:** Limpopo province (North-east) South Africa.  **Delivery mode (e.g., remotely online, in person):** - Data collected from records of two large public hospitals, South Africa Weather Service, and NOAA CPC.  **Intervention deliverers:**  **Timing and duration:**  **Intervention description:** No intervention | **Study type:**  Correlational study.  Wavelet transformation cross correlation analysis to monitor coincidences in changes of temp and rainfall and air quality variables with admissions to hospitals for GI illness, pneumonia, malaria and asthma.  **Review papers:**  **Length of follow-up:** N/A | **Outcome/s of interest:**  Associations between meteorological factors and hospital admissions, and estimated time lags between weather events and disease presentation. | **Main finding:**  New statistical estimate of time delay between change of weather and admissions for pneumonia and malaria  Increased pneumonia hospitalisation in lower temperatures, but also in hot/wet conditions.  Increased prevalence of pneumonia admissions 10-15 days after changes in air quality.  Increased malaria admissions 30 days following co-occurring rain and high temperature.  **Additional finding:**  **“**Causal relationships identified in this study between meteorological or air quality variables and health outcomes, together with specified time delay parameters between the onset of a driver and development of a disease, may be of use as parameter inputs to and non-trivial realistic tests for the predictive models and early warning systems of exposure and health impacts of climate change. They can additionally serve as a data-led understanding to inform relevant local actors and help facilitate novel adaptation measures in public health systems.” |
| **Author:** Ikeda *et al.*  **Year: 2017**  **DOI:**  [10.1038/s41598-017-02680-6](https://doi.org/10.1038/s41598-017-02680-6)  **Title:** Seasonally lagged effects of climatic factors on malaria incidence in South Africa  **Country:** South Africa  **Aim:** To analyse the relationship between both local climatic effects and remote atmospheric teleconnections on the incidence of malaria in Limpopo, including potential lag effects. | **Intervention:** No intervention    **Dates of data collection:** 1998-2014    **Population and sample size:** N/A    **Setting:** Limpopo, South Africa.    **Delivery mode (e.g., remotely online, in person):** -No intervention    **Intervention deliverers:** No intervention    **Timing and duration:** No intervention    **Intervention description:** No intervention | **Study type:** Descriptive cohort study    **Review papers:** N/A    **Length of follow-up:** N/A | **Outcome/s of interest:**     - El Nino - La Nina - Malaria incidence rate - Mean temperature - Precipitation | **Main finding:** A high incidence of malaria during the pre-peak season (Sep-Nov) was associated with the climate phenomenon La Niña and cool air temperatures over southern Africa. There was also high precipitation over neighbouring countries two to six months prior to malaria incidence. During the peak season (Dec-Feb). Warm temperatures and high precipitation in neighbouring countries were also observed two months prior to increased malaria incidence.    **Additional finding:**  This lagged association between regional climate and malaria incidence suggests that in areas at high risk for malaria, such as Limpopo, management plans should consider not only local climate patterns but those of neighbouring countries as well. These findings highlight the need to support cross-border control to minimize the spread of Malaria |
| **Author:** Ikeda *et al.*  **Year:** 2019  **DOI:**  [10.3390/atmos10090522](https://doi.org/10.3390/atmos10090522)  **Title:** Climatic factors in relation to diarrhoea hospital admissions in rural Limpopo, South Africa  **Country:** South Africa  **Aim:**  To explore the relationship between temperature, precipitation and diarrhoea case counts of hospital admissions among vulnerable communities living in a rural setting in South Africa | **Intervention:** No intervention  **Dates of data collection:**  January 2002 to December 2016  **Population and sample size:** All hospital admissions for diarrhoeal disease from two district hospitals. 11,228 cases in total over a 14 year period.  Daily precipitation and temperature data were obtained for the same period from the South African Weather Wervice.  **Setting:**  Limpopo province  **Delivery mode (e.g., remotely online, in person):** -  **Intervention deliverers:**  **Timing and duration:**  14 years data  **Intervention description:** No intervention | **Study type:**  Correlational study. Used contour analysis to visually explain observations in frequencies of high and low diarrhoea case counts in a season.  **Review papers:**  **Length of follow-up:** N/A | **Outcome/s of interest:**  Relationship between admissions for diarrhoeal disease in children under 5 and individuals over 5, and  precipitation and temperature | **Main finding:**  Children under 5 had the highest prevalence of diarrhoea during either wetter than average weather during the rainy season, or drier than average during the dry season, or other high temperatures.  In over 5s, higher rainfall was associated with lagged admissions for diarrhoeal disease. Rainfall likely to have a bigger role in diarrhoeal disease transmission than temperature, as lagged association was not seen for temperature related variables.  **Additional finding:**  The authors recommend the use of contour analysis as it is more suited to smaller data sets (missing data are common in handwritten rural hospital notes in SA) rather than time series analysis, as it focuses on anomalously high and low diarrhoea counts. . |

**Quality appraisals**

The Joanna Briggs quality appraisal tools for surveys and for qualitative studies were used for the following quality appraisals (Joanna Briggs Institute, 2017b, 2017c, 2017a)**.** See Tables 1-3.

**Table 1 JBI critical appraisal of systematic reviews**

| **Study** | **JBI Appraisal items** | | | | | | | | | | | **Score** |
| --- | --- | --- | --- | --- | --- | --- | --- | --- | --- | --- | --- | --- |
|  | **1** | **2** | **3** | **4** | **5** | **6** | **7** | **8** | **9** | **10** | **11** |  |
| Godsmark et al (2019) | Y | N | Y | U | N | N | N | Y | N | Y | Y | Moderate |
| Kapwata et al (2022) | Y | Y | U | U | N | N | N | Y | Y | Y | Y | Moderate |
| Khine and Langkulse (2023) | Y | Y | Y | Y | Y | Y | Y | Y | Y | Y | Y | High |
| Amegah et al., (2016) | Y | Y | Y | Y | Y | U | N | Y | N | Y | Y | Moderate |
| Chersich et al., (2018) | Y | Y | Y | Y | Y | N | U | U | N | Y | Y | Moderate |
| Wright *et al.* (2021) | Y | U | U | U | N | N | N | U | U | Y | Y | Low |

Key: Y – Yes; N – No; U – Unclear; n/a – not applicable

1. Is the review question clearly and explicitly stated?
2. Were the inclusion criteria appropriate for the review question?
3. Was the search strategy appropriate?
4. Were the sources and resources used to search for studies adequate?
5. Were the criteria for appraising studies appropriate?
6. Was critical appraisal conducted by two or more reviewers independently?
7. Were there methods to minimize errors in data extraction?
8. Were the methods used to combine studies appropriate?
9. Was the likelihood of publication bias assessed?
10. Were recommendations for policy and/or practice supported by the reported data?
11. Were the specific directives for new research appropriate?

**Table 2 JBI critical appraisal checklist for cross-sectional studies** (Joanna Briggs Institute, 2017a)

| **Study** | **JBI Appraisal items** | | | | | | | | **Score** |
| --- | --- | --- | --- | --- | --- | --- | --- | --- | --- |
|  | **1** | **2** | **3** | **4** | **5** | **6** | **7** | **8** |  |
| Colston *et al.* (2022) | Y | Y | U | Y | Y | Y | Y | Y | HIGH |
| Ikeda *et al.* (2017) | Y | Y | N | Y | Y | Y | Y | Y | High |
| Abdullahi, Nitschke and Sweijd (2022) | Y | Y | N | N | Y | Y | Y | Y | Medium |
| Ikeda *et al.* (2019) | Y | Y | Y | Y | Y | Y | Y | Y | High |
| Kapwata *et al.* (2021) | Y | Y | Y | Y | Y | Y | Y | Y | High |

Key: Y – Yes; N – No; U – Unclear; n/a – not applicable

1. Were the criteria for inclusion in the sample clearly defined?
2. Were the study subjects and the setting described in detail?
3. Was the exposure measured in a valid and reliable way?
4. Were objective, standard criteria used for measurement of the condition?
5. Were confounding factors identified?
6. Were strategies to deal with confounding factors stated?
7. Were the outcomes measured in a valid and reliable way?
8. Was appropriate statistical analysis used?

**Table 3 JBI critical appraisal scores for qualitative studies** (Joanna Briggs Institute, 2017b)

| **Study** | **JBI appraisal items** | | | | | | | | | | **Score** |
| --- | --- | --- | --- | --- | --- | --- | --- | --- | --- | --- | --- |
|  | **Q1** | **Q2** | **Q3** | **Q4** | **Q5** | **Q6** | **Q7** | **Q8** | **Q9** | **Q10** |  |
| Abrams et al (2021) | Y | Y | Y | Y | Y | U | U | Y | Y | Y | Moderate |
| Orievulu et al (2022) | Y | Y | Y | Y | Y | Y | Y | Y | Y | Y | High |

Key: CT: Can’t tell; N: No; Y: Yes

Q1: Is there congruity between the stated philosophical perspective and the research methodology?

Q2: Is there congruity between the research methodology and the research question or objectives?

Q3: Is there congruity between the research methodology and the methods used to collect data?

Q4: Is there congruity between the research methodology and the representation and analysis of data?

Q5: Is there congruity between the research methodology and the interpretation of results?

Q6: Is there a statement locating the researcher culturally or theoretically?

Q7: Is the influence of the researcher on the research, and vice- versa, addressed?

Q8: Are participants, and their voices, adequately represented?

Q9: Is the research ethical according to current criteria or, for recent studies, and is there evidence of ethical approval by an appropriate body?

Q10: Do the conclusions drawn in the research report flow from the analysis, or interpretation, of the data?

**Table 4 MMAT critical appraisal scores for mixed methods studies**

| **Study** | **MMAT**  **Qualitative items** | | | | | **MMAT Quantitative descriptive items** | | | | | **MMAT**  **Mixed methods items** | | | | | **Score** |
| --- | --- | --- | --- | --- | --- | --- | --- | --- | --- | --- | --- | --- | --- | --- | --- | --- |
|  | **1.1** | **1.2** | **1.3** | **1.4** | **1.5** | **4.1** | **4.2** | **4.3** | **4.4** | **4.5** | **5.1** | **5.2** | **5.3** | **5.4** | **5.5** |  |
| Lee *et al.* (2023) | Y | Y | Y | Y | Y | Y | Y | Y | Y | Y | Y | Y | Y | U | Y | High |

Key: Y – Yes; N – No; U – Unclear; n/a – not applicable

Qualitative:

1.1.	Is the qualitative approach appropriate to answer the research question?

1.2.	Are the qualitative data collection methods adequate to address the research question?

1.3.	Are the findings adequately derived from the data?

1.4.	Is the interpretation of results sufficiently substantiated by data?

1.5.	Is there coherence between qualitative data sources, collection, analysis and interpretation?

Quantitative Descriptive:

4.1.	Is the sampling strategy relevant to address the research question?

4.2.	Is the sample representative of the target population?

4.3.	Are the measurements appropriate?

4.4.	Is the risk of nonresponse bias low?

4.5.	Is the statistical analysis appropriate to answer the research question?

Mixed Methods:

5.1	Is there an adequate rationale for using a mixed methods design to address the research question?

5.2	Are the different components of the study effectively integrated to answer the research question

5.3	Are the outputs of the integration of qualitative and quantitative components adequately interpreted?

5.4	Are divergences and inconsistencies between quantitative and qualitative results adequately addressed

5.5	Do the different components of the study adhere to the quality criteria of each tradition of the methods involved?

**Reference:**

Munn, Z., Barker, T., Moola, S., Tufanaru, C., Stern, C., McArthur, A., Stephenson, M., Aromataris, E., 2021. Methodological quality of case series studies [WWW Document]. JBI Evid. Synth. URL [https://jbi.global/critical-appraisal-tools](https://eur01.safelinks.protection.outlook.com/?url=https%3A%2F%2Fjbi.global%2Fcritical-appraisal-tools&data=05%7C02%7CNatalie.Dickinson%40uws.ac.uk%7C2c47900fd9044b29b8ba08dce7b62568%7Cf89944b74a4e4ea791563299f3411647%7C0%7C0%7C638640017253638566%7CUnknown%7CTWFpbGZsb3d8eyJWIjoiMC4wLjAwMDAiLCJQIjoiV2luMzIiLCJBTiI6Ik1haWwiLCJXVCI6Mn0%3D%7C0%7C%7C%7C&sdata=q7AvN1mH34Q%2Boam%2FmJITg7sS6ww4ZsiCWgjgPalr8gI%3D&reserved=0) (accessed 6.30.21)

Ikeda, T., Behera, S. K., Morioka, Y., Minakawa, N., Hashizume, M., Tsuzuki, A., Maharaj, R., & Kruger, P. (2017). Seasonally lagged effects of climatic factors on malaria incidence in South Africa. Scientific Reports, 7(1). [https://doi.org/10.1038/s41598-017-02680-6](https://eur01.safelinks.protection.outlook.com/?url=https%3A%2F%2Fdoi.org%2F10.1038%2Fs41598-017-02680-6&data=05%7C02%7CNatalie.Dickinson%40uws.ac.uk%7C00f5e070cba24941549f08dcf282aeb3%7Cf89944b74a4e4ea791563299f3411647%7C0%7C0%7C638651892543845265%7CUnknown%7CTWFpbGZsb3d8eyJWIjoiMC4wLjAwMDAiLCJQIjoiV2luMzIiLCJBTiI6Ik1haWwiLCJXVCI6Mn0%3D%7C0%7C%7C%7C&sdata=dpytnfEoCFgPb%2FNqEj%2Fj4dYxLh%2FnXinIC6WE6foXO8o%3D&reserved=0)

Joanna Briggs Institute. (2017). Checklist for Analytical Cross Sectional Studies. Joanna Briggs Institute Reviewer’s Manual. [https://jbi.global/sites/default/files/2019-05/JBI_Critical_Appraisal-Checklist_for_Analytical_Cross_Sectional_Studies2017_0.pdf](https://eur01.safelinks.protection.outlook.com/?url=https%3A%2F%2Fjbi.global%2Fsites%2Fdefault%2Ffiles%2F2019-05%2FJBI_Critical_Appraisal-Checklist_for_Analytical_Cross_Sectional_Studies2017_0.pdf&data=05%7C02%7CNatalie.Dickinson%40uws.ac.uk%7C00f5e070cba24941549f08dcf282aeb3%7Cf89944b74a4e4ea791563299f3411647%7C0%7C0%7C638651892543874944%7CUnknown%7CTWFpbGZsb3d8eyJWIjoiMC4wLjAwMDAiLCJQIjoiV2luMzIiLCJBTiI6Ik1haWwiLCJXVCI6Mn0%3D%7C0%7C%7C%7C&sdata=1UTJQqMZA4%2Bou5oakzwGwZKpnu0lVaPVpRPj0X7st9E%3D&reserved=0)

### **References**

Godsmark, C. N., Irlam, J., van der Merwe, F., New, M., & Rother, H. A. (2019). Priority focus areas for a sub-national response to climate change and health: A South African provincial case study. In *Environment International* (Vol. 122, pp. 31–51). Elsevier Ltd. https://doi.org/10.1016/j.envint.2018.11.035

Joanna Briggs Institute. (2017a). *Checklist for Analytical Cross Sectional Studies*. Joanna Briggs Institute Reviewer’s Manual. https://jbi.global/sites/default/files/2019-05/JBI_Critical_Appraisal-Checklist_for_Analytical_Cross_Sectional_Studies2017_0.pdf

Joanna Briggs Institute. (2017b). Checklist for Qualitative Research. *Joanna Briggs Institute*, 6. http://www.joannabriggs.org/assets/docs/critical-appraisal-tools/JBI_Critical_Appraisal-Checklist_for_Qualitative_Research.pdf

Joanna Briggs Institute. (2017c). Checklist for Systematic Reviews and Research Syntheses. *Joanna Briggs Institute*. http://joannabriggs.org/research/critical-appraisal-tools.htmlwww.joannabriggs.org%0Awww.joannabriggs.org

Kapwata, T., Kunene, Z., Wernecke, B., Lange, S., Howard, G., Nijhawan, A., & Wright, C. Y. (2022). Applying a WASH Risk Assessment Tool in a Rural South African Setting to Identify Risks and Opportunities for Climate Resilient Communities. *International Journal of Environmental Research and Public Health*, *19*(5). https://doi.org/10.3390/ijerph19052664

Khine, M. M., & Langkulsen, U. (2023). The Implications of Climate Change on Health among Vulnerable Populations in South Africa: A Systematic Review. In *International Journal of Environmental Research and Public Health* (Vol. 20, Issue 4). MDPI. https://doi.org/10.3390/ijerph20043425
